# BrainXcan identifies brain features associated with behavioral and psychiatric traits using large scale genetic and imaging data

**DOI:** 10.1101/2021.06.01.21258159

**Authors:** Yanyu Liang, Owen Melia, Timothy J. Caroll, Thomas Brettin, Andrew Brown, Hae Kyung Im

**Affiliations:** Section of Genetic Medicine, University of Chicago, Chicago, Illinois, United States of America; Department of Computer Science, University of Chicago, Chicago, Illinois, United States of America; Department of Radiology, University of Chicago, Chicago, Illinois, United States of America; Computing Environment and Life Sciences Directorate, Argonne National Laboratory, Argonne, Illinois, United States of America; Consortium for Advanced Science and Engineering, University of Chicago, Chicago, Illinois, United States of America; Department of Population Health and Genomics, University of Dundee, Dundee, United Kingdom

## Abstract

Advances in brain MRI have enabled many discoveries in neuroscience. Comparison of brain MRI features between cases and controls have highlighted potential causes of psychiatric and behavioral disorders (complex traits). However, due to the cost of collecting MRI data and the difficulty in recruiting particular patient groups, most studies have small sample sizes, limiting their reliability. Furthermore, reverse causality complicates interpretation because many observed brain differences are the result rather than the cause of the disease. Here we propose a method (BrainXcan) that leverages the power of large-scale genome-wide association studies (GWAS) and reference brain MRI data to discover new mechanisms of disease etiology and validate existing ones. BrainXcan tests the association with genetic predictors of brain MRI-derived features and complex traits to pinpoint relevant region-specific and cross-brain features. As this approach requires only genetic data, BrainXcan allows us to test a host of hypotheses on mental illness, across many disorders and MRI modalities, using existing public data resources. For example, our method shows that reduced axonal density across the brain is associated with the risk of schizophrenia, consistent with the disconnectivity hypothesis. We also find structural features in the hippocampus, amygdala, and anterior cingulate cortex, among others associated with schizophrenia risk highlighting the potential of our approach, which uses orthogonal lines of evidence to inform the biology of complex traits.

## Introduction

Advances in MRI technology have enabled the measurement of brain structure, connectivity, and other features in a noninvasive manner. Many studies have successfully examined differences in brain features between diseased and healthy individuals to learn about the pathophysiology of brain-related diseases. However, the difficulty of recruiting patients and controls and the cost of the MRI scans have limited the sample sizes of these studies, greatly reducing their reproducibility (Marek et al., 2020).

On the other hand, genome-wide association studies (GWAS) use a very large number of participants (in some cases more than a million) to discover genetic mutations that affect brain-related diseases. For example, the Psychiatric Genomics Consortium (PGC) has performed GWAS of 11 psychiatric disorders, including attention-deficit-hyperactivity disorder, Alzheimer’s disease, autism, bipolar disorder, and schizophrenia, to elucidate their genetic basis. However, since these studies do not typically include brain imaging data, comparison of MRI images between cases and controls is not possible.

To bring together the advantages of the sophisticated and deep phenotypic data available in imaging studies and the very large sample sizes of GWAS studies, we turned to integrative approaches such as Transcriptome-wide association studies (TWAS). TWAS leverage GWAS studies and eQTL studies to genetically predict gene expression levels, test their correlation with complex traits and diseases, and ultimately, identify potentially causal genes (Gamazon et al., 2015; Gusev et al., 2016). Theoretically, this strategy could be applied to predict brain imaging phenotypes instead of gene expression data. However, the genetic variants regulating the expression of a given gene often have large effect sizes and are concentrated near the gene’s transcription start site (Wheeler et al., 2016). These properties limit the number of large-effect causal variants and make them easy to detect, simplifying prediction. In contrast, genetic predictors of brain features may be distributed throughout the genome, and their individual effect sizes are likely to be quite small, thus making them more difficult to optimize.

Here we propose BrainXcan, an extension of PrediXcan, that uses GWAS data (individual-level or summary statistics) to predict and compare image-derived brain features in cases and controls Gamazon et al. (2015). Rather than relying on a simple polygenic risk score to predict brain features (Knutson et al., 2020) or genetic correlations (Zhao et al., 2021), BrainXcan performs a comprehensive analysis of the genetic architecture to optimize prediction, which enhances both power and interpretability. Here, we apply BrainXcan to several complex traits using both large-scale GWAS summary statistics and UK Biobank’s individual-level data with close to half a million individuals. We demonstrate that BrainXcan can address the computational challenges and statistical issues stemming from the high polygenicity and genome-wide scale. Further, we show that BrainXcan can be used to corroborate existing hypotheses and generate new hypotheses about the role of the brain function and structure on psychiatric and neurological disorders.

## Results

### Overview of the BrainXcan framework

The BrainXcan framework is organized into three modules as outlined in Fig. 1. The **Prediction weight training module** trains linear genetic predictors of brain features, tests for association between brain features and genotype and calculates the sample covariance of the genotypes. These outcomes are saved for use in subsequent modules and shared publicly in `predictdb.org` with versioned and permanent records in zenodo.org.

**Figure 1:**
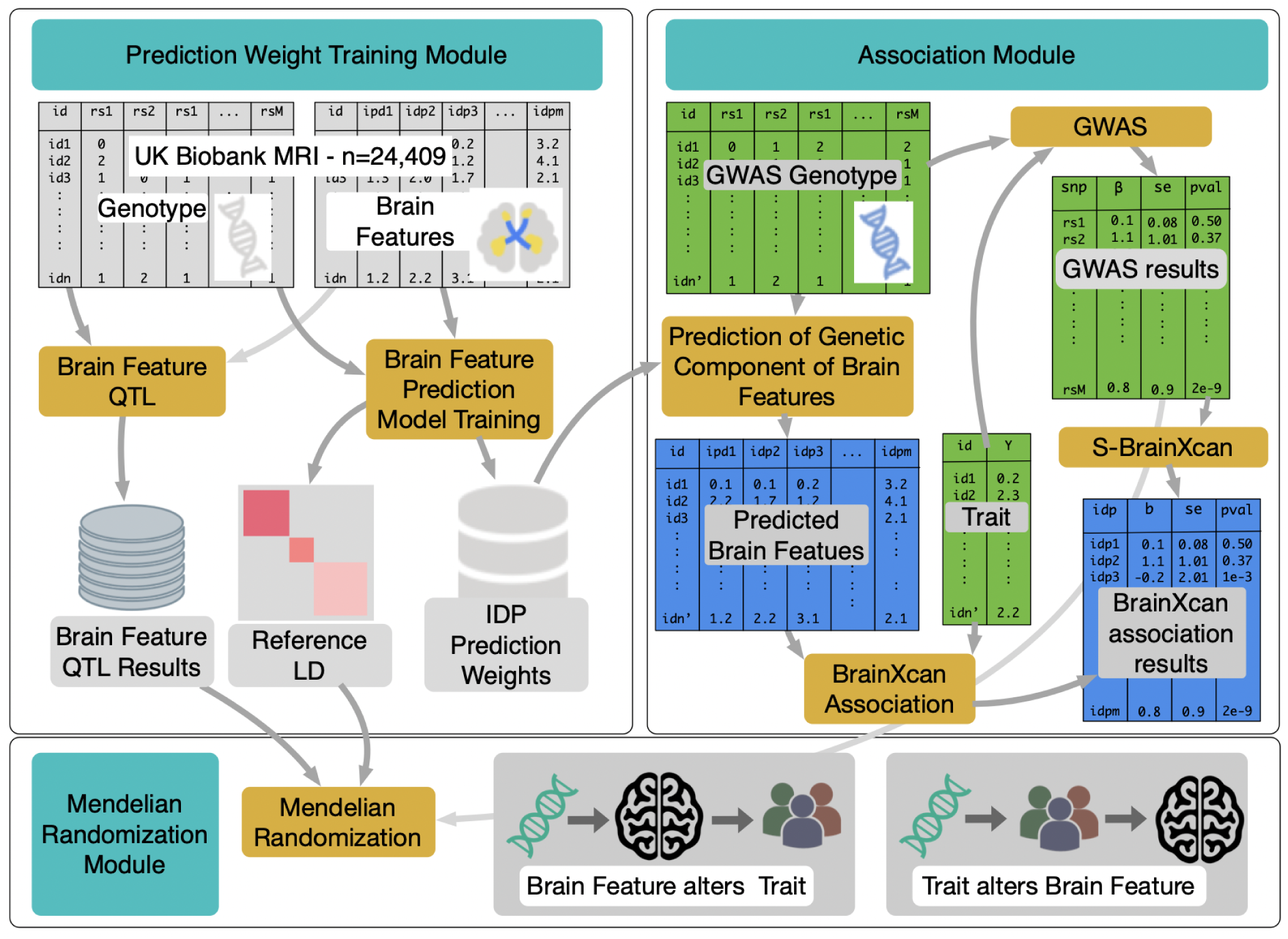
The workflow for implementing BrainXcan framework. The workflow is divided into three modules: Prediction Weight Training, Association, and Mendelian Randomization. The Prediction Weight Training module uses reference brain MRI data (containing brain features and genotype for a set of individuals) and trains genetic predictors of brain features (IDP Prediction Weights). It also computes the association between brain features and genotypes (Brain Feature QTL Results) and correlation between genotypes (Reference LD). This module only needs to be performed once and the results are provided to users as part of the BrainXcan software package. The Association and Mendelian randomization modules are the main analysis components that will be performed by most BrainXcan users. A typical user will have a trait of interest and the corresponding GWAS data. This input data (shown in green in the association module panel) can be in the individual-level format (GWAS genotype and Trait shown in green) or the summary (S-BrainXcan) format (GWAS Results shown in green). The Association module will generate a BrainXcan association results table (shown in blue). The significant brain features (with a p-value threshold defined by the user) will be processed with the Mendelian Randomization module to examine the direction of the putative causal flow and the consistency across multiple loci.

A typical user will not need to run the Training module, but will instead use the output that we have generated. They will have a trait of interest (typically a psychiatric, behavioral, or neurological disorder) with the corresponding GWAS data. The GWAS data can consist of a) the genotype and trait data (shown in green in Fig. 1) or b) the results table after performing the GWAS on the genotype and trait data.

The **Association module** will use the Brain Feature Prediction Weights and ‘Reference LD’ information we have shared to compute the BrainXcan association results table. The latter will provide estimated coefficients of the regression between the trait and the genetically predicted brain features, their standard error, and p-values.

When the individual-level data is provided, the Association module computes the Predicted brain features using the GWAS genotype and the corresponding prediction weights. Then, it calculates the association statistics between the predicted brain features and the trait of interest via linear regression, one feature at a time. For binary and other types of traits, logistic or other generalized linear models can be used. As usual, significant associations pinpoint candidate causal relationships between brain features and the trait. As explained in the brain feature processing section, we used features that represent both region-specific and brain-wide measurements.

The BrainXcan framework can still be applied to large-scale GWAS studies that do not have the individual-level data available. This is because the association statistics can be inferred using the summary results from GWAS, the brain feature prediction weights, and the reference LD data generated by the first module (see Methods). This approach was applied to genetic predictors of gene expression by Gusev et al. (2016) and Barbeira et al. (2018).

The **Mendelian randomization module** performs a number of multiple instrument-based Mendelian randomizations to determine the direction of the putative causal flow, i.e. whether alterations in brain features affect the complex trait or whether the trait (e.g. disease status) alters brain features. It provides bi-directional tests of causal flow and effect size scatter plots to help assess the consistency of the results.

### Prediction weight training module

#### Selection and preprocessing of brain MRI derived features

We downloaded uniformly preprocessed brain MRI image derived phenotypes (IDPs) from the UK Biobank. We selected 159 features derived from structural images representing total and gray matter volumes from different regions of the brain, and 300 diffusion MRI derived features representing neurite density, dispersion, and connectivity features. We focused on the 192 diffusion features mapped to the Johns Hopkins University’s 48-region atlas tracts (Wakana et al., 2004) After excluding related individuals and those of non-European ancestry, brain features on 24,409 individuals remained. We adjusted each feature for covariates including the first 10 genetic principal components (PCs), age at recruitment, sex, and four technical covariates indicating the location of the head in the scanner. See details in (Methods) and the list of brain features in table S1.

Moving forward, we processed the structural and diffusion MRI modalities separately. We categorized structural features into 5 subtypes: cortical gray matter volume, subcortical gray matter volume, subcortical total volume, cerebellum gray matter, and brain-stem, and included total gray matter volume and total gray and white matter volume as additional features. We also categorized diffusion MRI measures into 4 subtypes: fractional anisotropy (FA, a measure of diffusion along the white matter tracts), intracellular volume fraction (ICVF, an estimate of neurite/axonal density), isotropic volume fraction (ISOVF, an index of the relative extra-cellular water diffusion), and orientation diffusion index (OD, a measure of neurite dispersion) (Zhang et al., 2012).

For each subtype, we performed a principal components analysis. The first PC was a weighted average of features in the subtype suggesting that principal components could be used as proxies for the brain-wide features (fig. S1). For diffusion MRI subtypes, the first principal component explained 39% (FA), 53% (ICVF), 26% (ISOVF), and 20% (OD) of the total variance. For the structural MRIs, the first principal component explained 16% (cortical gray matter), 34% (subcortical gray matter), 37% (subcortical total volume), and 45% (cerebellum gray matter) of the total variability. As expected, after adjusting for the first principal component within each subtype, the correlation structure of the residual features was substantially reduced (figs. S2 and S3).

#### Generative models of brain features and complex traits

Since an important attribute of any method is the interpretability of the results, we sought to define brain features with the goal of facilitating interpretation. We prioritized the ability to tease out brain-wide effects from region-specific effects. i.e., determining whether a detected association with the trait was due to a feature that is common across the whole brain or specific to a region.

To distinguish between brain-wide effects and region-specific effects, we postulated the generative model shown in Fig. 2. The brain feature in each region (*F*_*k*_) is modeled as the sum of two independent latent components: a region-specific component (*R*_*k*_) and a brain-wide component (*L*). The observed value, IDP, is modeled as a noisy version of the region’s feature (IDP_*k*_ = *F*_*k*_ + *∈*_*k*_ = *L* + *R*_*k*_ + *∈*_*k*_). The parameter *s*^2^ determines the scale of the region-specific component (modeled as a normal random variable with variance *s*^2^) and *t*^2^ is the variance of the noise term *∈*_*k*_. Since we ultimately want to assess the significance of the association between predicted brain features and traits, multiplying the traits by a common scaling factor will not change the results. Accordingly, we chose the distribution of the latent variable to have variance equal to one.

**Figure 2:**
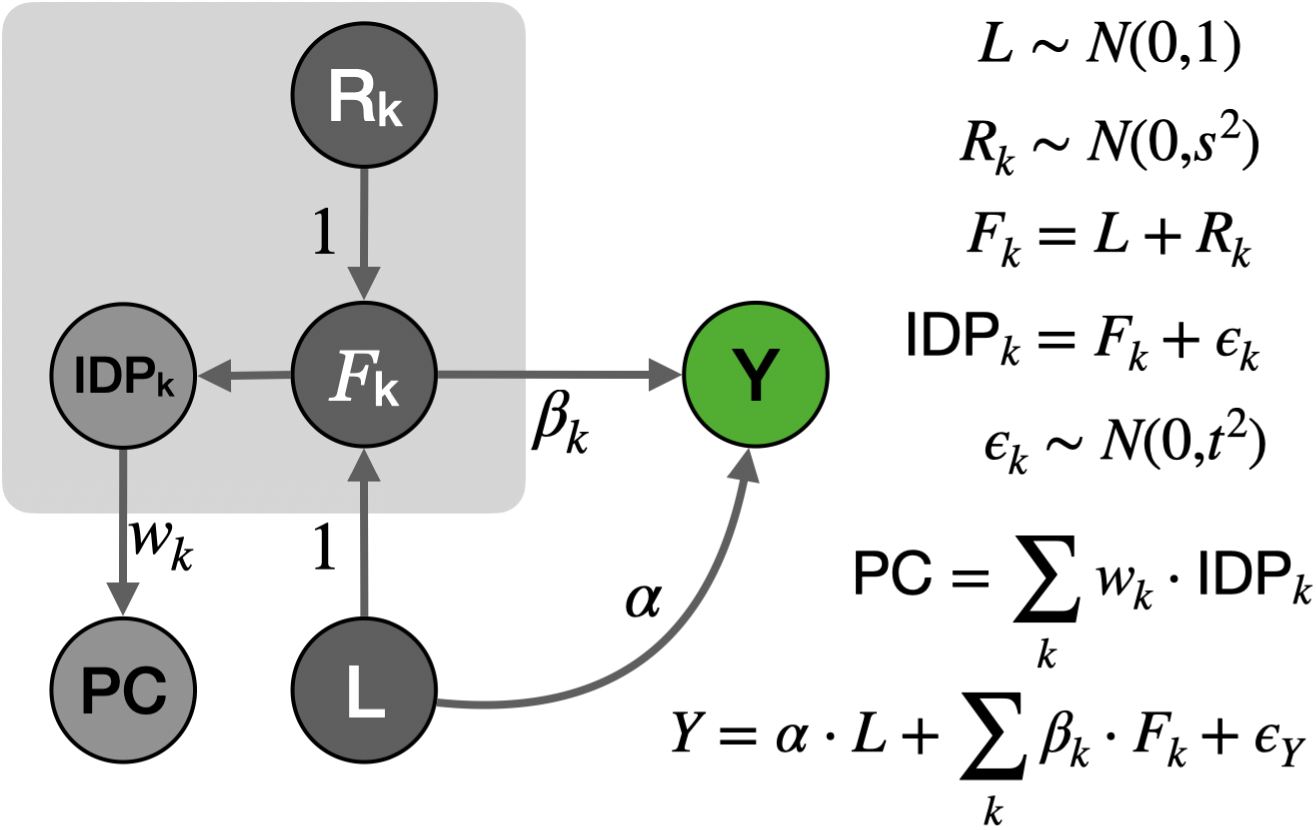
Generative model of brain features and complex traits. Brain features, *F*_*k*_, are modeled as a sum of a region-specific effect *R*_*k*_ and brain-wide component *L*. The observed brain features (IDPs) are considered to be noisy measurements of true features, *F*_*k*_. PCs of IDPs are weighted averages of IDPs. The complex trait *Y* is modeled as the sum of brain-wide effect (α) and region-specific effects (β_*k*_) and an error term (€_*Y*_).

Other approaches have incorporated additional principal PCs of raw brain features into their studies (Zhao et al., 2021). While we acknowledge that our decision to use only the first principal component was somewhat arbitrary, it simplifies the interpretability of our results and makes our method more user-friendly.

#### Genetic architecture of brain features

To determine whether brain features could be predicted using genetic data alone and to better inform optimal prediction approaches, we proceeded to investigate their genetic architecture. We calculated the heritability (roughly the magnitude of the genetic component) and degree of polygenicity (a measure of the number of independent genetic factors) of brain features.

**To calculate the heritability of brain features**, we used a standard mixed effects modeling approach (Yang et al., 2010) (Methods). Heritability estimates ranged from 5% to 43% with all the 95% confidence intervals above zero as shown in Fig. 3a. We note that the heritability of the principal components within each category (shown in red dots in Fig. 3a) is generally higher than the heritability of region-specific features (shown in black dots in Fig. 3a), providing support and validity to the principal components.

**Figure 3:**
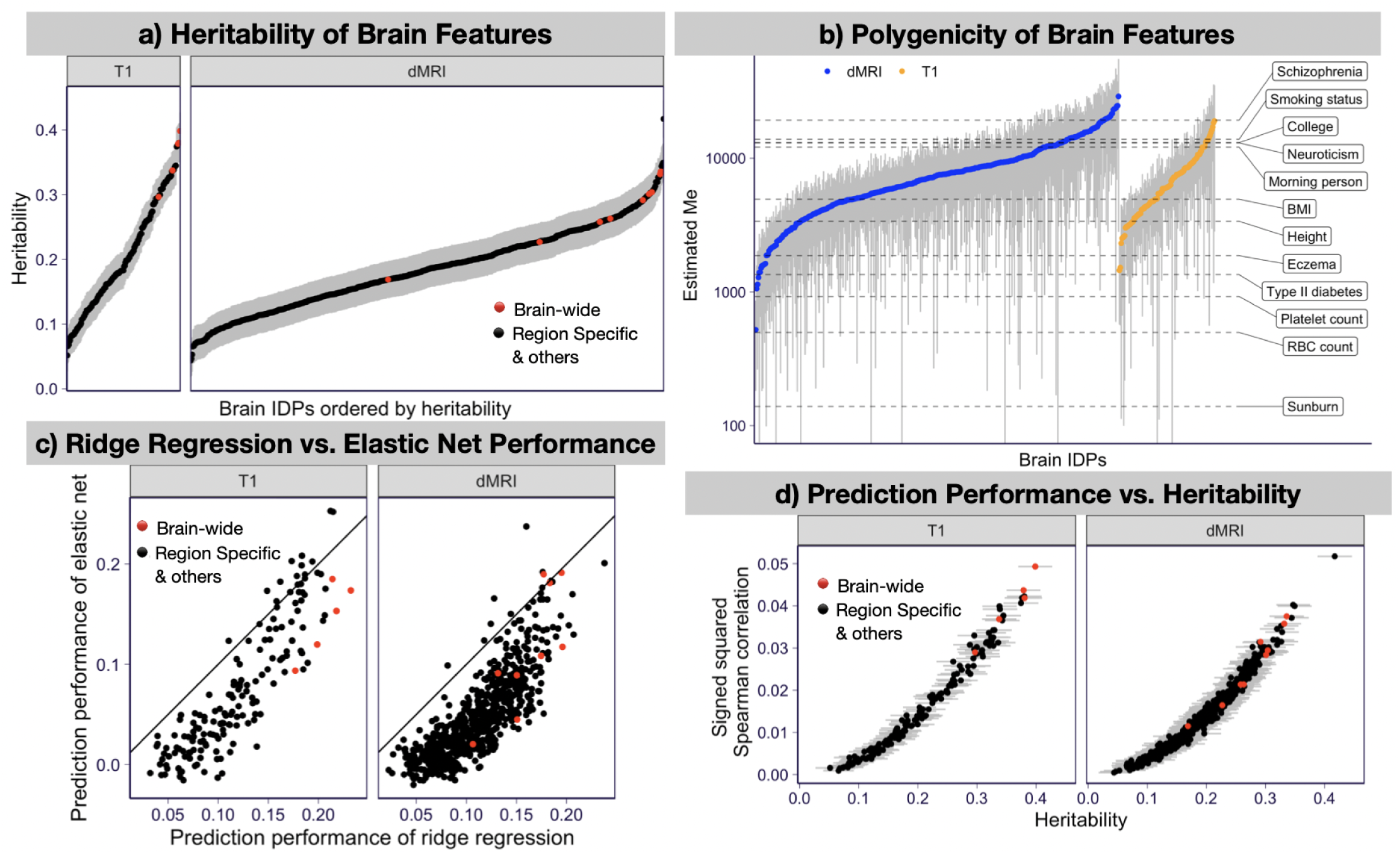
Genetic architecture and prediction performance of brain features. **a) Heritability of brain features**. Heritability and 95% confidence intervals are shown for structural (T1) and diffusion (dMRI) features. Red denotes common factors (PCs of each subtype) and black denotes the residual features in the subtype (after regressing out PCs). **b) Polygenicity of brain features**. The effective number of independently associated SNPs (*M*_*e*_) estimated using stratified LD fourth moments regression (O’Connor et al., 2019) for 522 brain features are shown on the y-axis. Gray bars indicate 95% confidence intervals. Horizontal dashed lines indicate the estimated *M*_*e*_ of 12 complex traits for reference. **c) Prediction performance of brain features**. This panel compares the performance of ridge regression vs. elastic net predictors measured by the Spearman correlation between the predicted and observed values in a five-fold cross-validated scheme. The performance of the ridge predictor is shown on the x-axis and the performance of the elastic net predictor is shown on the y-axis. The black solid line represents the identity line (*y* = *x*). The brain-wide features (IDP PCs) are shown in red and the remaining brain features are shown in black. **d) Prediction performance vs. heritability**. The signed squared Spearman correlations between observed and predicted IDPs of the ridge regression are shown on the y-axis and the estimated heritability are shown on the x-axis. The signed squared correlation is defined as sign(*x*) *· x* ^2^ for correlation *x* to preserve the sign of the correlation while taking the square. The error bar indicates the 95% confidence interval of the estimated heritability. The brain-wide features are in red and the rest of the brain features are in black.

**To quantify the degree of polygenicity of brain features**, we estimated the effective number of independently associated SNPs (*M*_*e*_) using stratified LD fourth moments regression (O’Connor et al., 2019) (Methods and fig. S4). Among 359 IDPs, 265 IDPs yielded a significant (*p <* 0.05) estimate of *M*_*e*_ with values ranging from 1,035 to 24,675, and a median of 6,245, which was higher than the estimates for canonical examples of polygenic traits such as height or BMI. The estimates are shown in Fig. 3b with common human traits added for reference.

These results indicate that the genetic prediction of brain features is feasible and suggest that predictors that include many genetic factors are likely to outperform sparser models with only a few genetic variants.

#### Ridge regression and elastic net prediction training

To maximize the applicability of our method, we designed BrainXcan to use only linear models. Linear predictors allow BrainXcan to be applied using only GWAS summary statistics - even when the individual-level genotype and trait data are not available. To keep the computation manageable, we restricted the training to a common set of SNPs in HapMap 3 (minor allele frequency, or MAF > 0.01 in European samples and MAF > 0.05 in UKB IDP cohort), a subset of SNPs that tends to be imputed with high quality across many existing GWAS. To avoid issues with strand misspecification, we excluded the ambiguous SNPs (e.g. AT, CG). These restrictions left us with a total of 1.07 million SNPs (Methods) for subsequent analyses.

We then used these data to train two sets of predictors, one with a ridge and the other one with an elastic net penalty. Ridge regression uses *l*_2_ penalty and yields highly polygenic predictors (all non-zero weights) whereas elastic net yields sparse models, setting the weights of most variants to 0. Given the high polygenicity of brain features, we expected ridge regression to perform better than the sparsity-inducing elastic net penalty (Methods).

To evaluate prediction performance, we calculated the Spearman correlation between the observed and predicted brain features with a five-fold cross-validation scheme (Methods), Fig. 3c. For our purposes, the correlation with the observed trait is a relevant measure of performance. We ultimately want to quantify the correlation between the brain feature and the complex trait, which is not impacted by a multiplicative factor.

For the ridge regression, the prediction performance ranged from 0.024 to 0.24 with a median of 0.13. For elastic net, the range was between -0.018 and 0.27 with a median of 0.075. All the ridge predictors and 95% of the elastic net predictors showed positive cross-validated Spearman correlations (fig. S7 and table S2), demonstrating the feasibility of genetically predicting brain features.

For most brain features, the ridge regression yielded higher cross-validated performance than elastic net, as we anticipated given the high polygenicity of the features (See Fig. 3c). To further corroborate our intuition that ridge regression performs better for more polygenic traits, we plotted the gain in performance by ridge regression over elastic net predictors against the estimated polygenicity. As hypothesized, we found that the gain increased with the degree of polygenicity of the brain feature (fig. S5).

Also, as expected, the prediction performance increased as the brain feature heritability increased (Fig. 3d). However, we also noted that the median prediction *R*^2^ was less than 8% of the heritability, an upper bound for the prediction. The low proportion of heritability captured by the genetic predictors highlights the need to increase the sample size of reference image data to reach the upper bound of the performance.

Moving forward, to reduce false positives in the BrainXcan association analysis, we filtered out unreliable predictors by keeping only brain features that showed prediction performance correlation greater than 0.1. Among structural features, 105 ridge predictors and 54 elastic net predictors passed the threshold from a total of 159 trained. Among 192 diffusion features, 148 ridge predictors and 62 elastic net predictors passed the threshold. All subtype-level PCs except the elastic net-based PC predictor of cortical region volumes were well predicted and retained for the subsequent analysis.

### Association module

#### Application of BrainXcan association module to complex traits

We selected 9 traits from the UK Biobank and performed BrainXcan association using 327,743 individuals of British ancestry. As described in the overview section, we calculated the genetically predicted brain features for all individuals and correlated them with the traits using linear regression. The traits included alcohol consumption, smoking, coffee consumption, depression, parental depression, parental Alzheimer’s disease, handedness, BMI, and height (see detailed list on table S2). To avoid overfitting issues, we excluded individuals used to the train of the prediction models.

#### S-BrainXcan uses GWAS summary statistics to compute associations

Next, we extended the BrainXcan association module so that it could infer the association statistics from the GWAS summary statistics without the use of individual-level genotype and trait data. We term this version S-BrainXcan. Our approach is similar to the S-PrediXcan method and other summary-based TWAS approaches used for correlating genetically predicted transcriptomes with complex traits (Gusev et al., 2016; Barbeira et al., 2018). However, unlike gene expression prediction, which only uses variants in the vicinity of the gene, IDP prediction requires a much larger number of genetic predictors distributed throughout the genome. To make IDP prediction computationally feasible, we developed a scalable method that can handle this added complexity (Methods). Specifically, we calculated the covariances between the SNPs used in the prediction and saved them in a sparse format, setting the correlations between distant SNPs (i.e. separated by more than 200 SNPs) to be 0. We chose this number because it represents a trade-off between the accuracy of the covariances and the size of the reference files needed by the users. We decided to use the HapMap3 subset of SNPs for the prediction in anticipation of this computational burden.

We applied S-BrainXcan association analysis on 35 traits for which we did not have access to the individual-level data but had GWAS results. These traits included behavioral, psychiatric and neurologic traits, height, and body mass index (see table S3).

For standing height and BMI traits, we confirmed the reliability of the S-BrainXcan by comparing the z-scores of the associations to the ones obtained from individual-level BrainXcan using data from the UK Biobank. Overall, z-scores were highly concordant reassuring us that the approximations used in the implementation of the summary version of BrainXcan did not have a detrimental effect on the results (fig. S6).

**To assess the calibration of the type 1 error** (false positive rate), we tested for association between the brain feature and the trait under the null, where both the trait and the brain feature were heritable but not related to each other. We also calculated the association between the trait and the brain feature calculated with block permuted prediction weights.

Somewhat unexpectedly, we found that the z-scores of the association between brain features and traits were inflated (i.e., they were not normally distributed with mean 0 and variance 1). To correct for the inflation, we divided the raw BrainXcan z-scores by the standard deviation of the z-scores under the permuted null. (Methods). We added this correction to the BrainXcan software to ensure proper type 1 error calibration of the results.

**To assess the sensitivity of BrainXcan to the prediction approach**, we compared the BrainXcan z-scores obtained by using ridge vs elastic net-based predictors. Reassuringly, we found high concordance between ridge and elastic net results. Ridge predictors yielded higher significance, as expected given their increased prediction performance relative to elastic net (fig. S7). **We focus on ridge-based results for the remainder**.

Combining both summary level and UK Biobank traits, 90% of tested brain features (236 out of the 261) were significantly associated with at least one trait. As expected, better powered GWAS traits with a larger number of significant associations also yielded more significant feature-to-trait associations.

Brain-wide features represented tended to be more significantly associated, suggesting a more prominent role in complex traits compared to region specific features (fig. S8). However, the increased significance could be also due to the fact that they tended to be better predicted and hence less affected by attenuation bias (Supplementary Notes 1).

To verify replication of our results, we compared BrainXcan z-scores from independent studies for height, neuroticism, depression, BMI, intelligence, and Alzheimer’s disease. Reassuringly, we found highly concordant z-scores between the independent studies (Fig. 4a).

**Figure 4:**
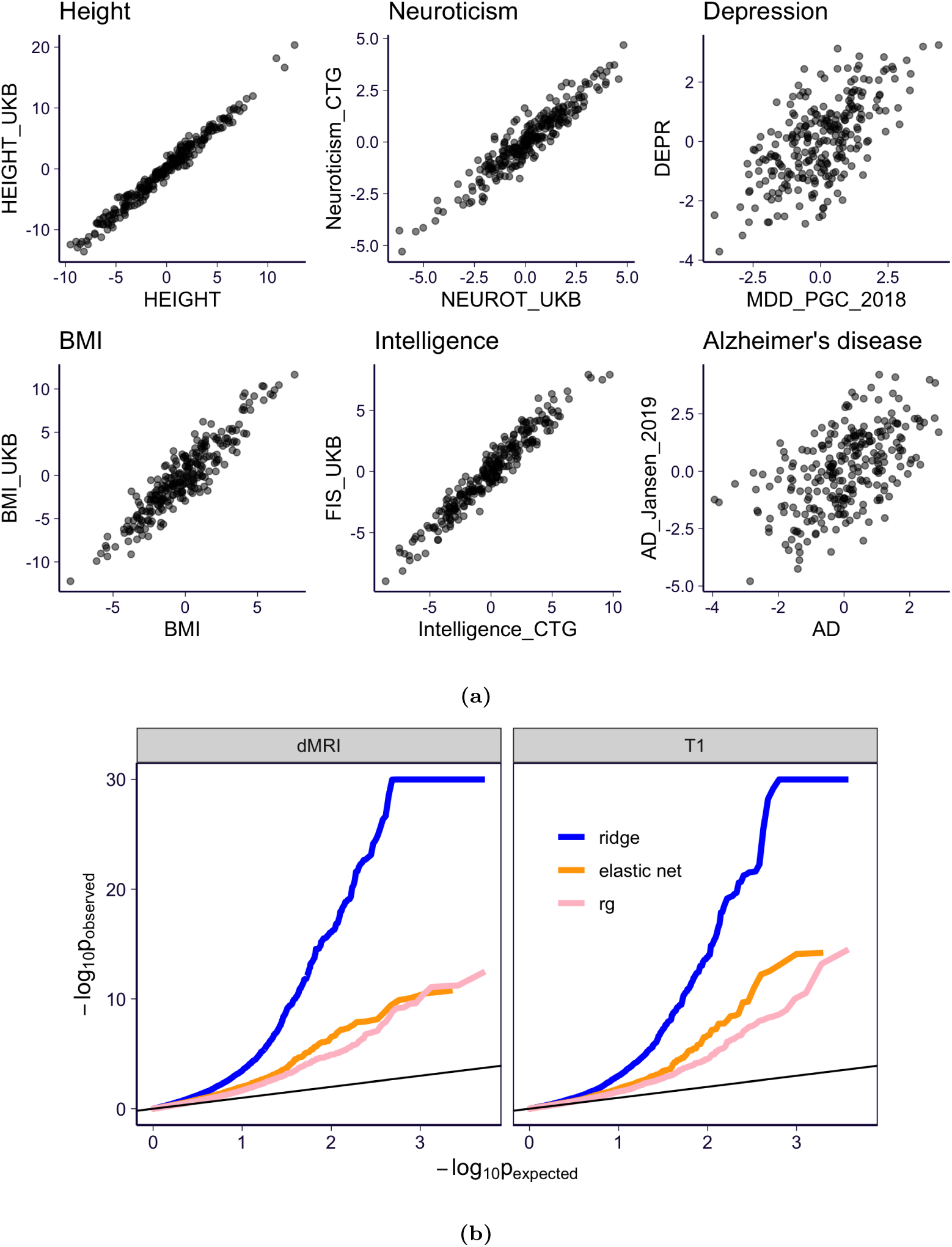
S-BrainXcan association statistics for 35 GWAS. **(a)** Six traits which have multiple GWAS were analyzed by S-BrainXcan are shown. For each brain feature, the S-BrainXcan z-scores (from ridge models) from the two GWAS are shown on x-axis and y-axis respectively (see the GWAS label in table S3). **(b)** The S-BrainXcan p-values are shown as a QQ-plot (against the expected p-values under the null). For visualization purposes, the observed p-values smaller than 1 *×* 10^*−*30^ are set to 1 *×* 10^*−*30^. Label ‘rg’ represents the genetic correlation result. The black lines are the identity line (*y* = *x*).

#### Genetic correlations yield similar but less significant associations

Genetic correlations between brain features and complex traits can provide, in principle, similar information to BrainXcan association results. We computed the correlations for all feature-trait pairs and compared the results to BrainXcan associations. (Methods ; Fig. 4b). We found that the z-scores of the genetic correlations were highly correlated to the z-scores of BrainXcan associations (with correlations ranging from 0.51 to 0.97, with a median of 0.81; fig. S9). However, BrainXcan yielded 4-fold larger number of significant feature/trait associations compared to the genetic correlation approach suggesting that using optimal predictors of brain features yield more power to identify putative causal links.

### Mendelian Randomization module

Mendelian randomization evaluates the causal relationship between traits 1 and 2 by testing whether increased “exposure” to the first trait (represented as the trait 1 GWAS effect size) is associated with an increased or decreased level of the second trait (indicated by the trait 2 GWAS effect size). In Mendelian randomization settings, individuals are thought to be randomized at meiosis to either inherit the risk-increasing allele or not. Because of this parallel to randomized trials, the level of causal evidence derived from Mendelian randomization is considered to be higher than that of observational studies, but lower than that of actual randomized trials (the gold standard for causal determination used in clinical trials).

It is possible to infer the direction of the causal flow by selecting genetic variants that have strong effects on the first trait and testing for a significant association with the effect sizes of the second trait and vice versa. As described below, scatter plots of effect sizes showing the two selection strategies (by significance of trait 1 or 2), are added to the automated reports.

We applied several Mendelian randomization approaches, including MR-BASE (Hemani et al., 2018), inverse variance weighted regression (Burgess et al., 2013), weighted median method (Bowden et al., 2016), and Egger regression (Bowden et al., 2015), to determine the direction of the putative causal flow, i.e. whether changes in IDPs are affecting changes in GWAS traits. In all analyses, we assumed that multiple genetic variants are strongly associated with the phenotype (we refer to these as variants as “instruments”). For complex traits, we selected SNPs with GWAS p-values *<* 5 *×* 10^*−*8^ and for IDPs, we selected SNPs with GWAS p-values *<* 10^*−*5^. The less stringent p-value for IDPs was necessary to have a sufficient number of instruments, which can be adjusted as the number of individuals in the IDP dataset increases. To streamline the interpretations of the multiple Mendelian randomization results, we combined the p-values of each Mendelian randomization output using an extension of the ACAT method (Liu et al., 2019), which accounts for concordance of the sign of the results. Results for all Mendelian Randomization approaches are provided with the output of the software. See caveats in interpreting the Mendelian Randomization results in Supplementary Notes 4 and fig. S10.

### Application of BrainXcan to schizophrenia risk

To demonstrate its features, we applied the full BrainXcan pipeline to a schizophrenia GWAS Ripke et al. (2020). We focused on 261 features, which include 48 cortical gray matter volumes, 10 subcortical volumes, 13 subcortical gray matter volumes, fractional anisotropy (water diffusivity along nerve tracts) in 46 regions, ICVF (intracellular volume fraction) in 44 regions, OD (orientation dispersion) in 45 regions, and ISOVF (isotropic volume fraction) in 13 regions (note that features in the ISOVF category were less heritable leading to fewer successful predictors). We also included brain-wide measures represented by leading principal components of each subtype (gray volumes of cortical, cerebellum, and subcortical regions, subcortical total volumes, FA, ICVF, OD, and ISOVF).

Among the 261 features, 2 were significantly associated with risk of schizophrenia after Bonferroni correction (p-value < 0.05/261). Figs. 7 and 8 provide a snapshot for schizophrenia risk. To aid interpretation, an interactive annotation of different regions of the brain is added to the output of the software’s automated pipeline (See example in https://brainxcan.hakyimlab.org/post/2021/05/06/brainxcan-automated-reports/#interactive-annotation-of-regions).

Among diffusion MRI associations, the principal component of ICVF, a proxy for brain-wide axonal density, was the most significant association for schizophrenia risk, with lower axonal density associated with higher risk of schizophrenia (Fig. 5). The principal component of fractional anisotropy, a brain-wide measure of water diffusion efficiency along nerve tracts, was also negatively associated with schizophrenia, while the OD index (dispersion of neurite orientation along tracts) showed no significant association with schizophrenia.

**Figure 5:**
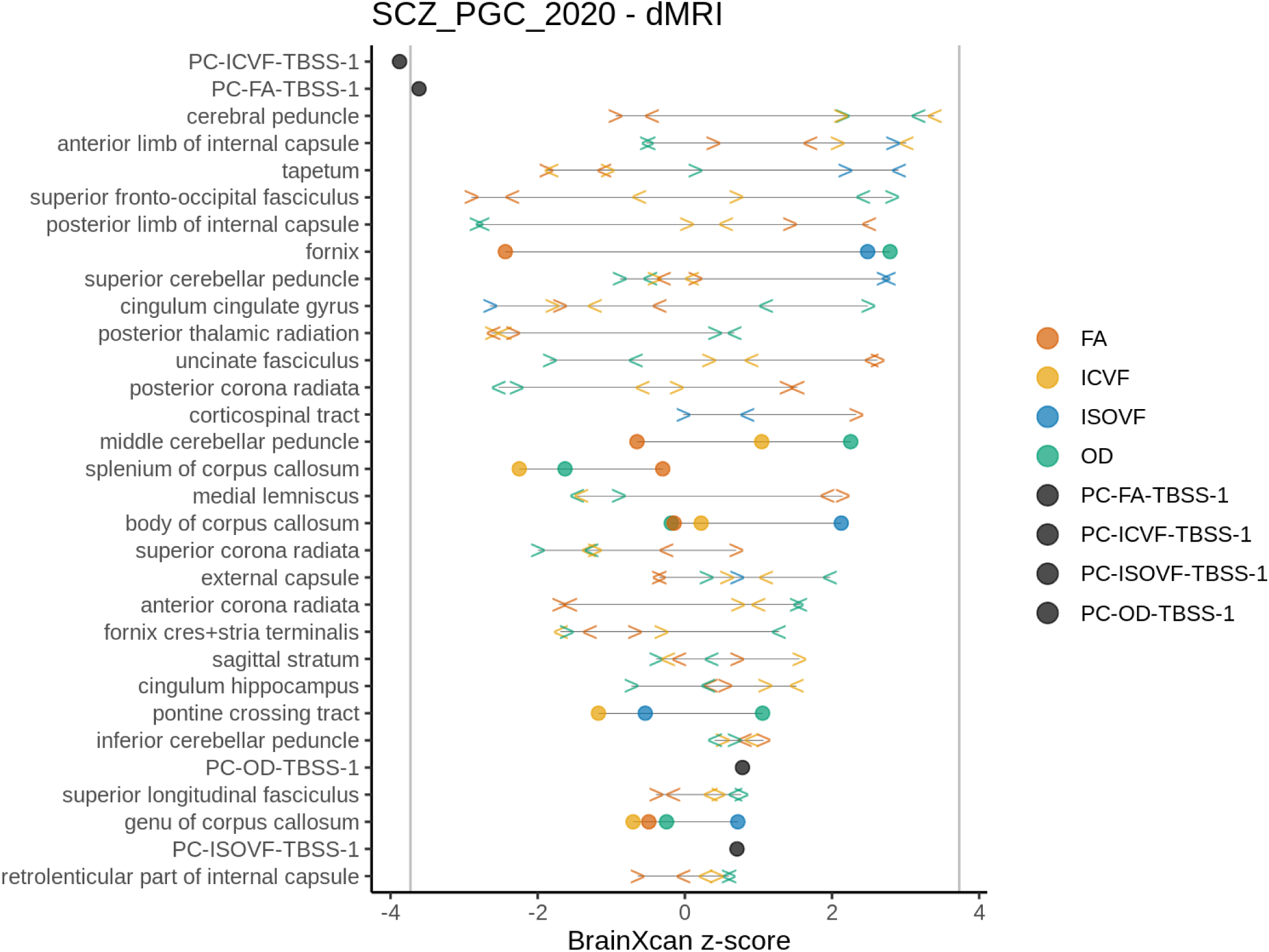
Schizophrenia risk association with diffusion MRI. Z-scores of region-specific feature associations with schizophrenia risk using GWAS effect sizes reported in Ripke et al. (2020). Features starting with “PC” correspond to brain-wide properties (principal components of IDPs). FA: fractional anisotropy, ICVF: intracellular volume fraction, ISOVF: isotropic volume fraction, OD: orientation dispersion index. *<* indicates left, *>* indicates right, circles are used when sides are not defined

The total volume of the hippocampus (right side, relative to brain size) was positively associated with schizophrenia risk, as were grey matter volumes of the frontal orbital complex, the anterior cingulate cortex, and posterior temporal fusiform cortex. In contrast, the total volume of the thalamus (both sides, relative to brain size) was negatively associated with schizophrenia risk. (Fig. 6). Observed hippocampus, thalamus, amygdala, anterior cingulate cortex volumes have all been reported to be associated with schizophrenia status (van Erp et al., 2016; Shepherd et al., 2012).

**Figure 6:**
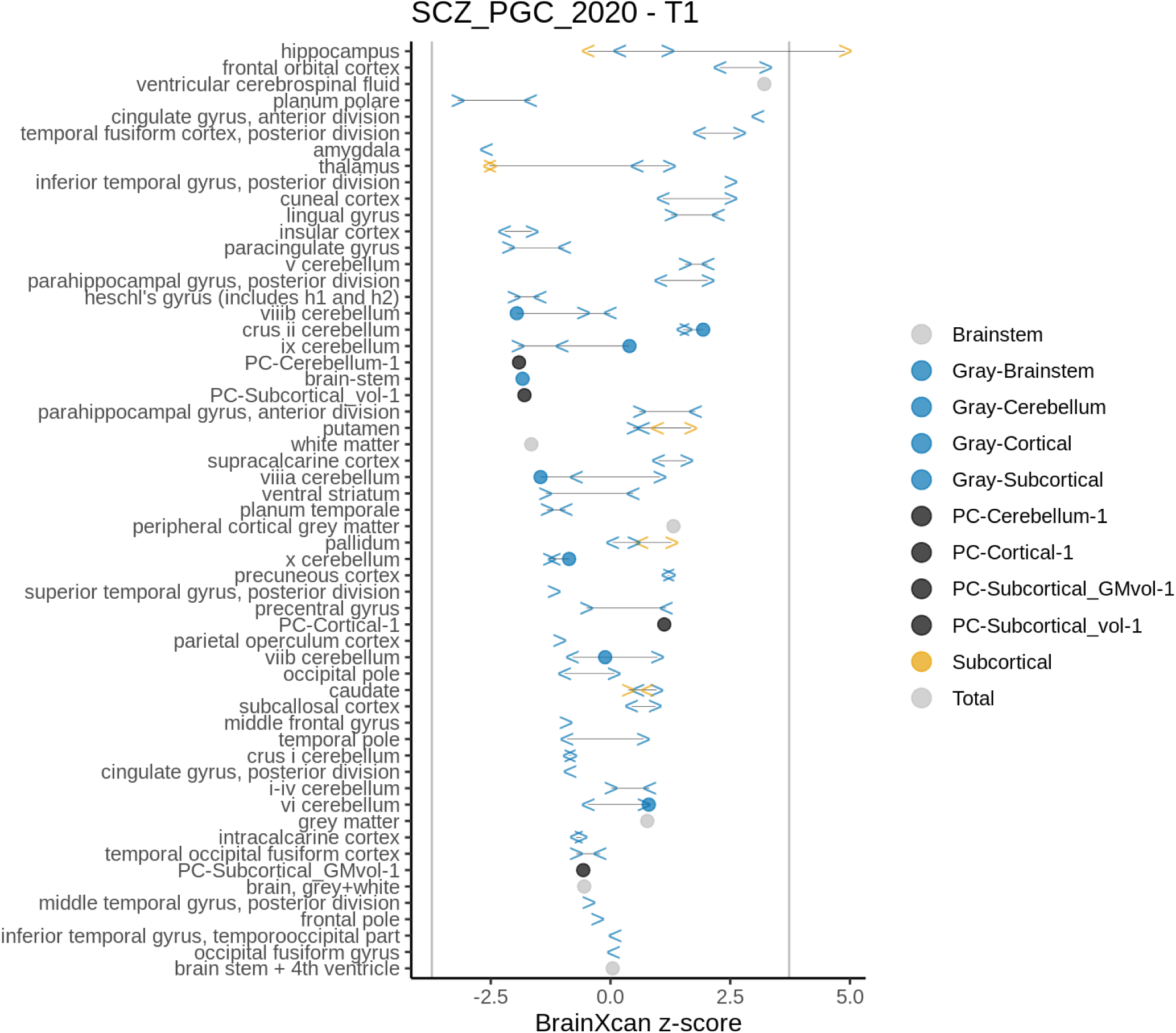
Schizophrenia risk association with structural features. Z-scores of region-specific feature association with schizophrenia risk using GWAS effect sizes reported in Ripke et al. (2020). Features starting with “PC” correspond to brain-wide properties (principal components of IDPs). In blue are shown the gray matter volumes of the cortex, subcortex, brainstem, and cerebellum. In black are shown the principal components of each category. In yellow are shown the associations with subcortical total volumes quantified with FIRST. *<* indicates left, *>* indicates right, circles are used when sides are not defined.

**Figure 7:**
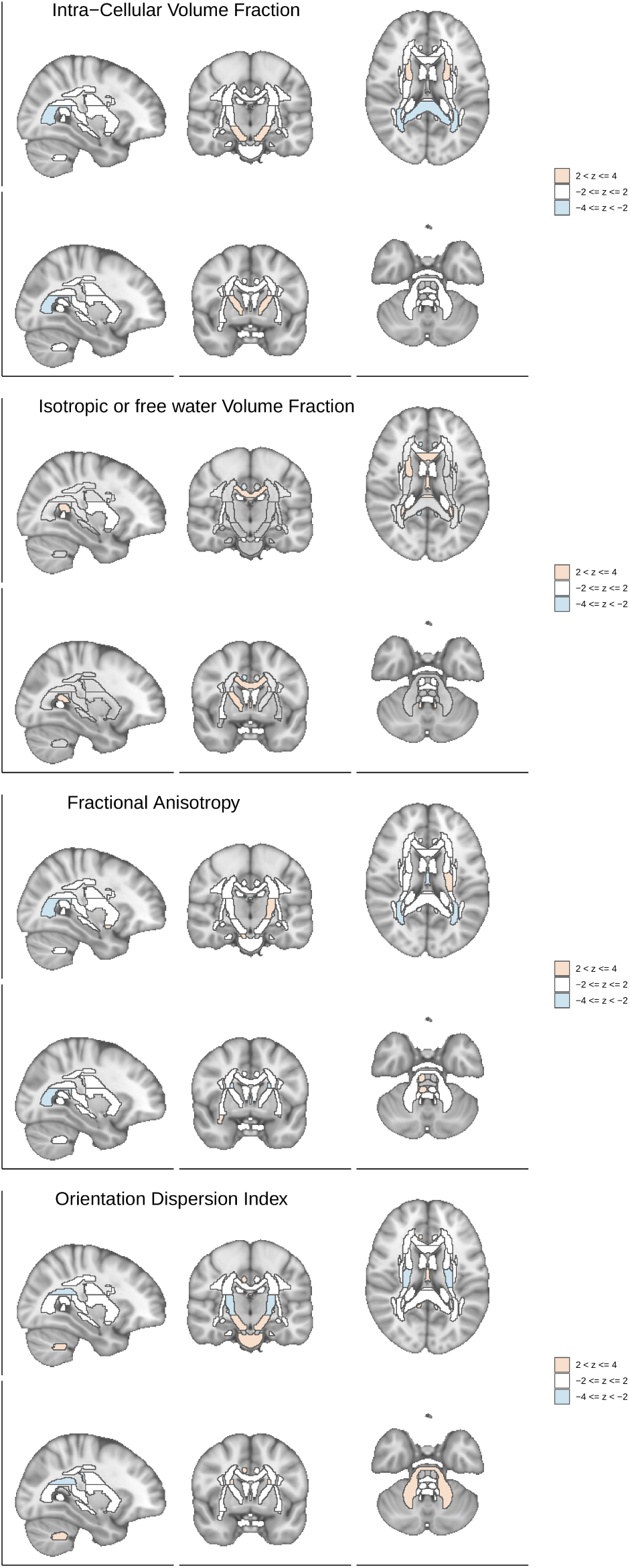
Brain visualization of diffusion features associations with schizophrenia risk. Z-scores of the associations between the brain region and schizophrenia risk are shown with different slices of the brain.

**Figure 8:**
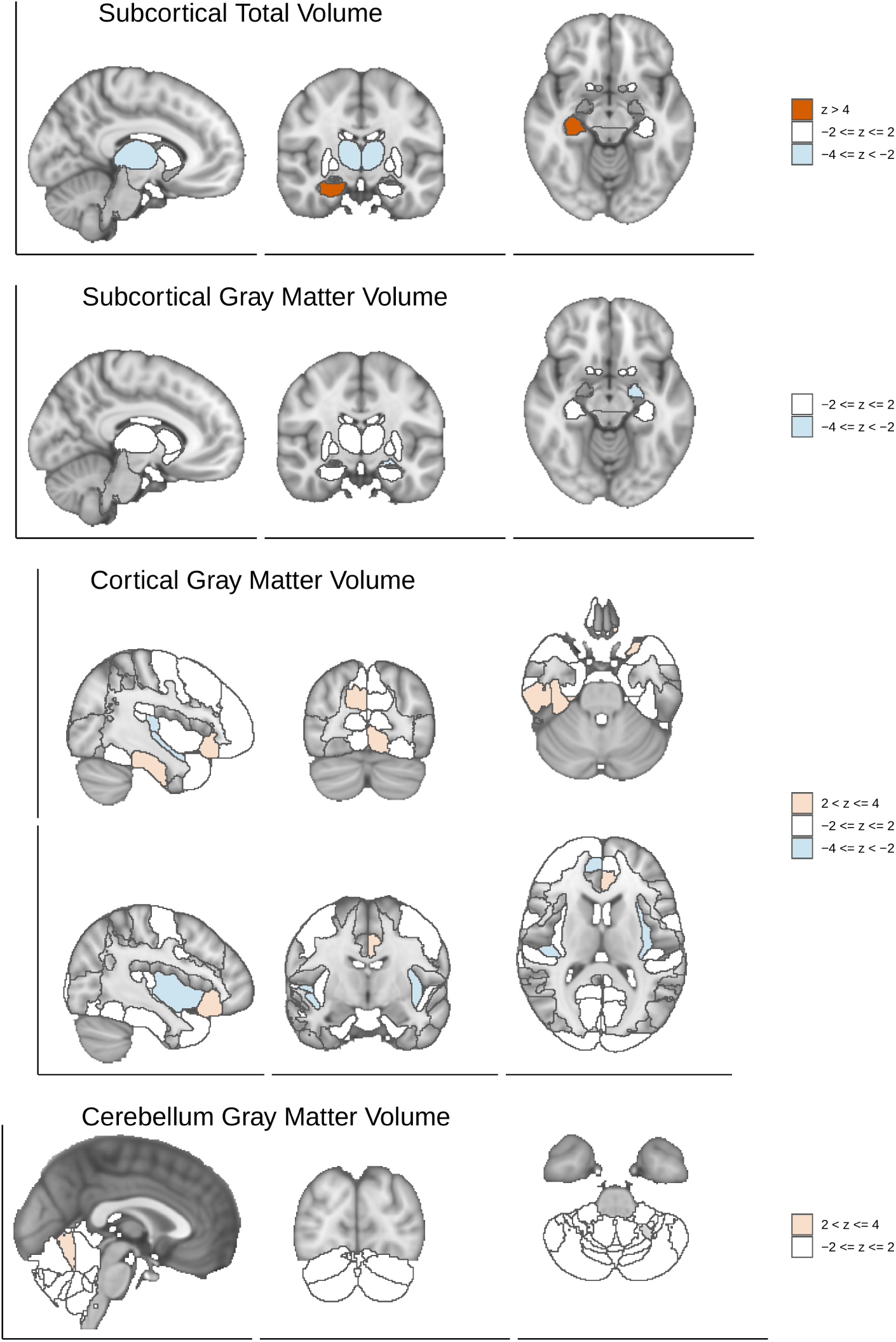
Brain visualization of structural features associations with schizophrenia risk. Z-scores of the associations between the brain region and schizophrenia risk are shown with different slices of the brain.

Among the top 10 features associated with schizophrenia, we found two instances of brain feature to schizophrenia risk causal flow with nominally significant p-value (p*<* 0.05) and five instances in the opposite direction. Notice that our reference image data did not include individuals with schizophrenia. Therefore, our design cannot detect schizophrenia to brain feature effects (scenario ii) in fig. S10).

## Discussion

We propose a robust and scalable framework we call BrainXcan, which leverages genetically predicted brain features trained in reference MRI datasets, GWAS of complex diseases, and computational and methodological advances to dissect the biology of behavioral, neurological, and psychiatric traits. Our approach addresses the sample size limitations of MRI studies by taking advantage of increasing cohorts of GWAS studies and large MRI data in predominantly healthy subjects. The use of genetic variation helps us circumvent the reverse causality problem.

Our association module identifies brain features likely to influence behavioral and psychiatric traits as well as other features that can be modified by the disease. Our Mendelian randomization module quantifies the evidence for both directions of the putative causal flow (brain feature to disease or vice-versa). Naturally, both directions of the effects are informative. Understanding how human disease causes changes in brain features captured by MRI can help design better diagnostic tools. Brain features that modulate disease risk provide insights into disease pathogenesis and can help identify preventive or therapeutic strategies.

To encourage broad adoption of BrainXcan by users less familiar with genetic tools, we provide a user friendly pipeline and an automated report. All the tools necessary to perform prediction, association, and causal flow assessment is provided (https://github.com/hakyimlab/brainxcan).

In the process of developing BrainXcan, we learned that, similar to psychiatric and behavioral traits, brain features are highly polygenicin many cases even more so than human height (the canonical example of a polygenic trait). Their shared similarity in genetic architecture suggests that brain features can be useful endo-phenotypes to improve the classification of complex psychiatric diseases.

We present an application to schizophrenia to showcase the potential of our method.

The significant association between low levels of brain-wide ICVF (a proxy for axonal density) with high risk of schizophrenia corroborates the long standing hypothesis that schizophrenia is a disorder of disconnectivity. They are also consistent with reduced FA in schizophrenia cases compared to controls as reported by Kelly et al. (2018). However, since our technique uses healthy individuals for MRI-based genetic prediction of brain features, they are less likely to capture a consequence of the disease.

We also find the volumes of many regions of relevance associated with schizophrenia risk, including the amygdala, hippocampus, and the anterior cingulate cortex. Our results add robust orthogonal lines of evidence to existing literature since our associations are based on the genetic components of the traits and are less likely to be confounded than studies with observed traits.

We note that there are several limitations in the current work. First, the prediction performance of the current genetic predictors are largely limited by the size of the training cohort (Fig. 3d). As the UK Biobank is gradually collecting more brain imaging data (Littlejohns et al., 2020), we expect that the training cohort size will increase to at least 100,000 individuals within the next few years. We will be updating the brain feature predictors as more data becomesx available. Second, the S-BrainXcan calculation relies on the genotype covariance, which is approximated as a banded matrix (Methods). This approximation may affect the stability of the joint analysis, since the joint analysis relies heavily on the predicted feature covariance, which is derived from the genotype covariance. This was one of the reasons we decided not to pursue the joint model, since joint models can be more sensitive to LD-misspecification and hence lead to false positives. Third, the BrainXcan analysis cannot establish the causal relationship between the brain features and the complex trait. Although we run Mendelian randomization in both the forward and the reverse directions for the feature/trait candidate, the results should be interpreted with caution due to the following reasons: i) different Mendelian randomization tests may not give consistently significant results; ii) the Mendelian randomization of the forward and reverse directions typically have different power due to differences in sample sizes and genetic architecture; iii) if the same GWAS is used for both BrainXcan association and Mendelian randomization, the Mendelian randomization p-value is not well-calibrated; iv) causality is valid only when the Mendelian randomization assumptions are held. Given these limitations, the Mendelian randomization results should not be interpreted as definitive support for the direction of the causal flow. Increased reference image datasets and additional model development that account for these limitations are needed. Moreover, only linear prediction models are used in our implementation. Although more sophisticated models could be used for prediction, this would limit the application to cases where the full individual level data is available. However, given the difficulties of accessing and handling individual level data at a very large scale, a linear model seems reasonable. Despite these limitations, we anticipate that BrainXcan, a user-friendly analysis tool, will be broadly adopted and help in identifying brain features important in the pathogenesis as well as diagnosis of complex traits.

## Methods

### Preprocessing of UK Biobank brain features

We queried the UK Biobank database using ukbREST (Pividori and Im, 2019) to retrieve the list of 459 MRI image-derived phenotypes (IDPs, also referred to as brain features or features) as shown in table S1 (Smith et al., 2020). Among the features, 300 were derived from diffusion MRI and 159 from T1-weighted structural measurements. For the diffusion features, we focused our analysis on the 192 features mapped to the Johns Hopkins University’s 48 region atlas tracts (Wakana et al., 2004). The T1-weighted structural measurements include 139 FAST-based grey matter segmentation-based measurements and 14 FIRST-based subcortical structures measurements and 6 T1 structural brain MRIs measuring the total volumes of the peripheral cortical grey matter, ventricular cerebrospinal fluid, brain grey matter, brain white matter, brain grey and white matter, and brain stem. See technical details in Miller et al. (2016). In total, we collected 24,409 European-descent individuals in UK Biobank with non-missing brain features and non-missing values for other covariates, such as genetic PCs, sex, and age at recruitment.

We scaled the structural features using the volumetric scaling factor from the T1 head image (UK Biobank Data-Field 25000) so that the measurement of the brain region volume was relative to the total brain volume. Additionally, we regressed out the following scanner position covariates: UK Biobank data fields 25756 (Scanner lateral (X) brain position), 25757 (Scanner transverse (Y) brain position), 25758 (Scanner longitudinal (Z) brain position), and 25759 (Scanner table position). We also regressed out the first 10 genetic PCs, age, sex, age squared, age *×* sex, and age squared *×* sex.

To adjust for the correlation between features and to extract the factor in common across different regions of the brain, we performed principal component analysis on the IDP matrices (individual-by-IDP matrices) within each IDP subtype. For T1 IDPs, the subtypes were gray matter volume of the cortical regions, gray matter volume of the subcortical regions, total volume of subcortical regions, and gray matter of the cerebellum regions. For diffusion IDPs, the subtypes were defined by the four measure types (FA, ICVF, ISOVF, and OD). We consider the first PC to represent the common factor for each IDP subtype. To define the region-specific components, we calculated the residuals after regressing out the first PC from the IDPs (see discussion in Supplementary Note 2). To improve robustness and reduce the influence of outliers, we inverse-normalized the PCs and the residuals.

### Selecting genetic variants from UK Biobank imputed genotypes

To reduce the computational burden, we restricted the prediction to the HapMap 3 subset of variants with minor allele frequency above 1% among CEU (European-descent) individuals (International HapMap 3 Consortium and others, 2010). Among these variants, we kept variants with MAF *>* 5% in UKB IDP cohort (all of European-descent). We also excluded ambiguous variants which have reverse complementary bases as the reference and the alternative alleles (AT and CG pairs). In total, 1,078,323 variants passed our criteria and among these, 1,071,650 variants were present in the UK Biobank imputed genotype data (by SNP rs ID). In the subsequent analysis, we limited the computation to this set of variants.

### Estimating the heritability

We estimated the heritability of IDPs assuming random effects for SNPs (Yang et al., 2010). We used the EMMA algorithm proposed in Kang et al. (2008) to avoid repeated calculations when dealing with multiple phenotypes from the same cohort. In the analysis, the genetic relatedness matrix (GRM) was built using the pre-selected HapMap 3 SNPs. Since we regressed out MRI technical covariates, the first 10 genetic PCs, age, sex, squared age, age *×* sex, and squared age *×* sex in the pre-processing step, we only added the intercept as a covariate in the heritability calculation.

### Estimating of polygenicity

To quantify polygenicity, we estimated the effective number of independently associated SNPs using the stratified LD fourth moments regression method (O’Connor et al., 2019). We downloaded the pre-computed LD scores and LD fourth moments from https://www.dropbox.com/sh/iiyftw01gdpt6un/AACU7AmWK45RxTmDJvRkdKhIa?dl=0 and used the scores stored in `baselineLD.1kg.l2l4.mat`. These scores were based on baselineLD annotations (Gazal et al., 2017). The stratified LD fourth moments regression was performed in MATLAB by calling `SLD4M` function shared in https://github.com/lukejoconnor/SLD4M (O’Connor et al., 2019). To obtain the effective number of independently associated common SNPs, we aggregated the estimated *M*_*e*_ values across 10 MAF bins which correspond to the common variants (MAF *>* 0.05). The aggregation was done by setting `report_annot_mat` variable in the `SLD4M` function.

### Building polygenic predictors of brain features

We built elastic net and ridge regression predictors of brain features, one feature at a time. As mentioned above, the prediction used the pre-selected 1,078,323 common HapMap3 variants. Since we already adjusted for covariates, we included only the intercept as a covariate in the model training.

### Ridge regression models

For ridge regression we perform the following optimization problem:

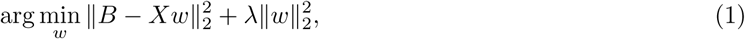

where *B* is *n ×* 1 the mean-centered brain feature (fitting o ne a t a t ime), *X* i s t he *n × P* standardized genotype matrix (mean centered and divided by the standard deviation) of the variants, and *w* is the *P ×* 1 prediction weights. Since the number of variants (*P*) is much larger than the number of samples (*n*), instead of using the usual ridge regression solution

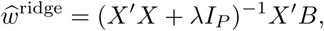

we use the equivalent form

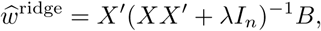

which can be derived using the ‘matrix push through identity’. *λ* is a hyperparameter to be determined. *I*_*P*_ and *I*_*n*_ are *P × P* and *n × n* identity matrices.

Let Σ represent the GRM matrix and we have S = *XX ′/P*, where *X* is the genotype matrix in the training set. The expression for the *w*^ridge^ can be re-parameterized with 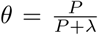 which takes value in the 0 to 1 range.

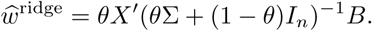

We chose the optimal hyperparameter *θ* by performing 5-fold cross-validation on a grid of *θ* values 0.01, 0.05, 0.1, 0.2, 0.3, 0.4, 0.5, 0.6, 0.7, 0.8, 0.9, 0.95.

#### Elastic net models

We trained the elastic net predictors using the R package `snpnet` which implements the BSAIL algorithm proposed by Qian et al. (2020). Specifically, we fi t th e fo llowing op timization problem:

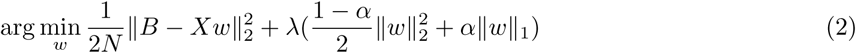

We set the mixing hyperparameter *α* to 0.1. To choose the optimal hyperparameter *λ*, we divided the brain reference data into a training set with 80% of the samples and a validation set with the remaining 20% of the samples. We performed elastic net regression using a grid of *λ* values determined by `snpnet` (default parameters were used) and selected the *λ* that yielded the largest correlation between predicted and actual *B* in the validation set. Finally, we trained the final prediction model using the full set of data (training and validation). When using `snpnet`, we set the maximum number of iterations (`niter`) to 100 and the number of SNPs to consider in each batch (`num.snps.batch`) to 200. Each brain feature was trained separately.

#### Calculating the prediction performance

For both ridge and elastic net models, we evaluated the prediction performance by 5-fold cross validation. For each fold, we used the remaining 4 folds to train the prediction models using the procedures described above. The prediction weights were used to predict on the held-out fold of the data. This procedure was repeated for all five splits and the predicted values were aggregated across all folds. The prediction accuracy was evaluated in terms of *R*^2^, Pearson correlation, and Spearman correlation.

### BrainXcan with summary statistics

When the individual-level information is not available, we calculated the BrainXcan statistic approximately using the GWAS summary statistic and the genotype covariance from a reference panel.

The formulas are similar to Gusev et al. (2016) and Barbeira et al. (2018). Briefly, let 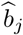 and 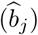 be the estimated effect size and the corresponding standard error for variant *j* from the GWAS. And *z*_GWAS,*j*_ is the GWAS z-score of SNP *j*. Let 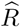 represent the genotype sample covariance matrix where 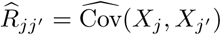, namely the sample covariance between variant *j* and *j*^*/*^. We can calculate the marginal test statistics using the following results:

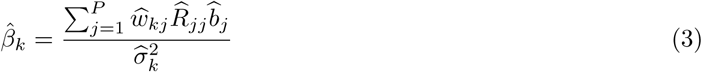

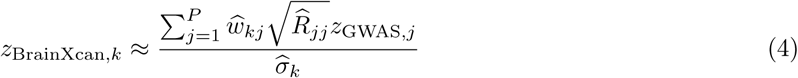

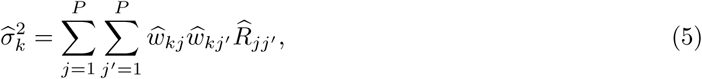

where *z*_BrainXcan,*k*_ represents the BrainXcan marginal test z-score for the *k*th brain IDP. We refer this summary statistic-based BrainXcan as S-BrainXcan.

In principle, we need to consider the genotype covariance for all the genome-wide variant pairs. To reduce the computational burden, we first assumed that the between-chromosome covariance is zero. Moreover, we considered the per-chromosome genotype covariance matrix as a banded matrix with bandwidth equal to 200. In other words, any variant pairs with more than 200 variants in-between are considered to have zero covariance. Results remain substantively similar when using larger number of SNPs, up to 1000. As shown in the results, the concordance between summary- and individual-level based results reassured us that this was a reasonable trade off while keeping the total size of covariance files that need to be shared with the user below 50GB. We used the set of 24,409 UK Biobank individuals included in the brain IDP model training as the reference panel for genotype covariance calculation.

### Adjusting BrainXcan z-scores

To account for the potential inflation of BrainXcan z-scores, we performed a permutation procedure to obtain the BrainXcan z-score under the permutation-based null. We permuted the variant labels in the prediction weights so that, for each variant, potential dependencies among brain IDP weights are preserved in the permutation. To keep the potential local LD pattern in prediction weights, the permutation was done at the LD block level, i.e. variants in the same LD block remain in relative order in the permutation. We used LD blocks derived from European individuals (Berisa and Pickrell, 2016). We performed the permutation 10 times and calculated BrainXcan z-scores using the permuted weights, which resulted in 10 *×* #*{*IDP*}* z-scores under the permutation-based null. We adjusted the BrainXcan z-scores by dividing the standard deviation of the z-scores under the permutation-based null by its value, namely

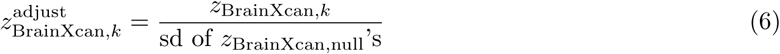

### Performing GWAS for brain features

We performed GWAS for all the brain IDPs using the Python package `tensorqtl` (Taylor-Weiner et al., 2019). Since we regressed out MRI technical covariates, the first 10 genetic PCs, age, sex, squared age, age *×* sex, and squared age *×* sex in the pre-processing step, the intercept was the only covariate included in the `tensorqtl` runs.

### Mendelian randomization analysis of IDP/phenotype pairs

To investigate the direction of the effect, we performed Mendelian randomization analysis for the significant feature/trait pairs identified in the BrainXcan association stage. The MR analysis was performed for both directions: i) brain feature *→* complex trait, treating the brain feature as the mediating phenotype and the trait of interest as the outcome trait; ii) trait *→* brain feature, the trait of interest as the mediating phenotype and the brain feature as the outcome trait. As the inputs of the analysis, we used the brain IDP GWAS results as described above. For the phenotype of interest, we used the GWAS results which were also used for the S-BrainXcan analysis. The MR analysis was performed using the R package `TwoSampleMR` (Hemani et al., 2018).

For the mediating trait, the instrument variants were selected using the LD clumping function (`ld_clump`) in the R package `ieugwasr` (Elsworth et al., 2020). We used the EUR super-population in 1000 Genomes data (1000 Genomes Project Consortium, 2015) as the LD reference panel, which we downloaded from http://fileserve.mrcieu.ac.uk/ld/1kg.v3.tgz. The LD clumping parameters were `clump_kb = 10000` and `clump_r2 = 0.001`. The p-value parameter (`clump_p`) in the LD clumping was 10^*−*5^ for IDP GWAS and 5 *×* 10^*−*8^ for phenotype GWAS, which gave approximately independent and significant variant instruments.

We reported the MR results using three MR methods: i) inverse variance weighted MR (Burgess et al., 2013); ii) median-based estimator: weighted median (Bowden et al., 2016); iii) MR Egger analysis (Bowden et al., 2015), which corresponds to `mr_ivw`, `mr_weighted_median`, and `mr_egger_regression` in `TwoSampleMR`. We further reported a meta-analyzed p-value summarizing the results of the three MR tests being performed. The meta-analysis is based on an extension of ACAT method (Liu et al., 2019) that takes into account the direction of the effects. See Supplementary Notes 3 and fig. A2 for additional details.

### Calculating the genetic correlation for brain feature/trait pairs

The genetic correlation between a brain feature and the trait of interest was calculated using the cross-trait LD Score regression (Bulik-Sullivan et al., 2015) implemented in the Python package `ldsc` (https://github.com/bulik/ldsc). The pre-computed LD-scores were downloaded from https://storage.googleapis.com/broad-alkesgroup-public/LDSCORE/eur_w_ld_chr.tar.bz2 which are based on the 1000 Genomes European data. We used the brain feature GWAS results as described above and the GWAS data for each phenotype was the same as the data used for the S-BrainXcan analysis.

## Data Availability

Data is available in https://github.com/hakyimlab/brainxcan and links therein

## Acknowledgements

This research has been conducted using the UK Biobank Resource under Application Number 19526. YL, OM, and HKI received partial funding from P30DK020595, U01HG009086. We thank Natalia Gonzales, Desiree Leach, and Beau Burnett for help editing the paper.

## Code and data availability

https://github.com/hakyimlab/brainxcan

## Contributions

Y.L. designed and built the computational pipelines, performed the data analysis, created the software and visualization for BrainXcan, and wrote the original draft of the manuscript. O.M. performed initial data cleaning and exploration and contributed to the pipeline development. T.J.C. discussed and interpreted the results. T.B. provided the computational support and discussed and interpreted the results. A.B. discussed and interpreted the results. H.K.I. supervised the whole project, designed the computational pipelines and data analysis, and extensively edited the manuscript. All authors read, edited, and approved the final manuscript.

