## Supplementary Material for "BrainXcan identifies brain features associated with behavioral and psychiatric traits using large scale genetic and imaging data"

### Supplementary Materials: BrainXcan: Associating genetically predicted image-derived phenotypes with complex traits to inform their biology

Yanyu Liang[1,*] Owen Melia[2] Thomas Brettin[3,4] Andrew Brown[5] Hae Kyung Im[1,*]

**1** Section of Genetic Medicine, University of Chicago, Chicago, Illinois, United States of America
**2** Department of Computer Science, University of Chicago, Chicago, Illinois, United States of America
**3** Computing Environment and Life Sciences Directorate, Argonne National Laboratory, Argonne, Illinois, United States of America
**4** Consortium for Advanced Science and Engineering, University of Chicago, Chicago, Illinois, United States of America
**5** Department of Population Health and Genomics, University of Dundee, Dundee, United Kingdom

### Contents of Supplementary Notes

**1 Deriving bias of BrainXcan estimates** **13**

**2 Using IDP residual instead of fitting IDP and PC jointly** **15**

**3 Aggregating Mendelian Randomization test results by extending the Aggregated Cauchy Association test (ACAT) method** **18**

**4 Caveats on interpreting Mendelian randomization results** **21**

### Supplementary Figures

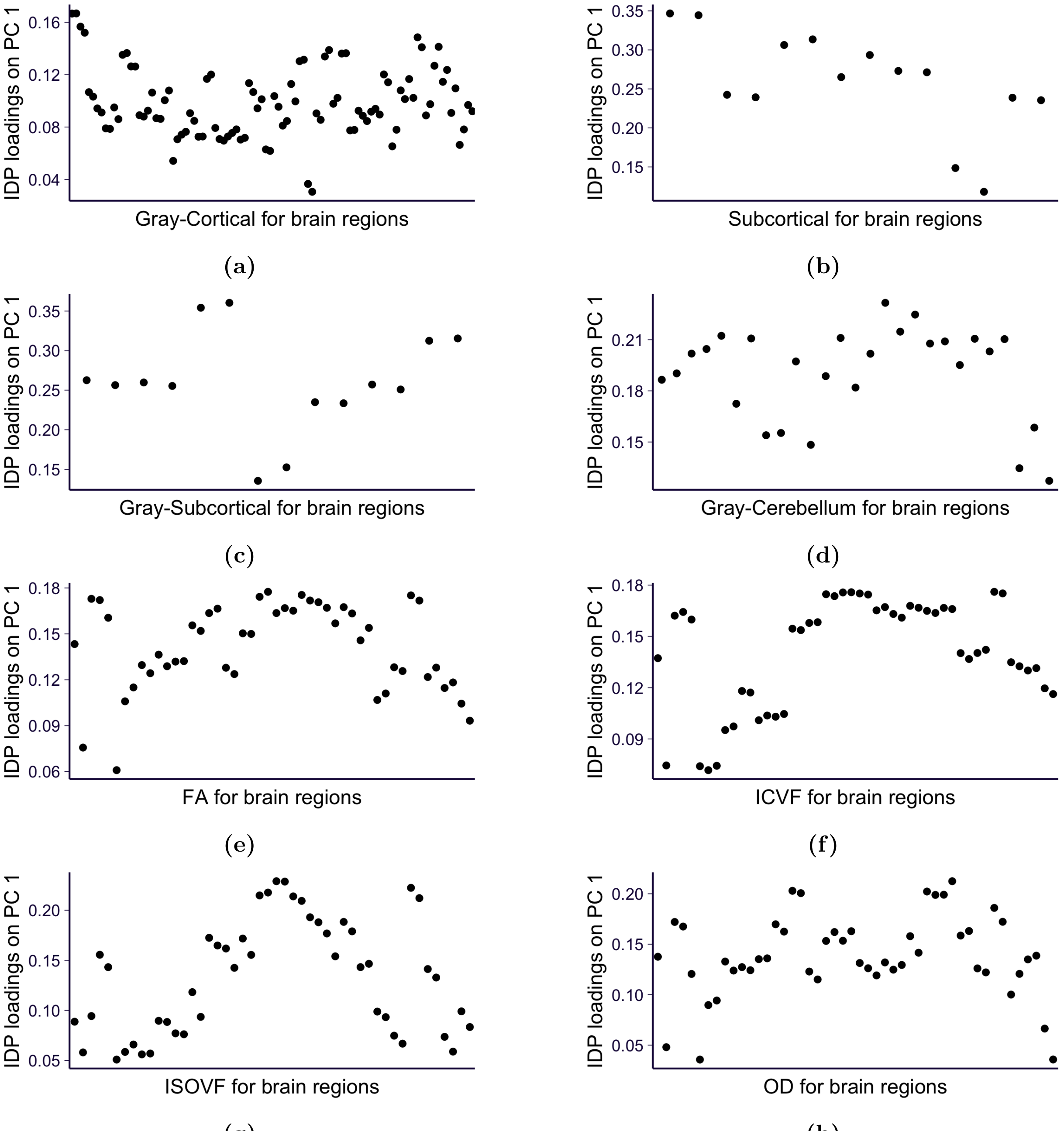

**fig. S1. The first PC of each IDP modality** For each IDP modality, the contribution of PC1 to each of the brain IDPs within the modality group is shown. Panel **a)** to **d)** show results for T1 modalities: gray matter volume of cortical regions, total volume of subcortical regions, gray matter volume of subcortical regions, and gray matter volume of cerebellum regions. Panel **e)** to **h)** show results for TBSS-based dMRI modalities: FA, ICVF, ISOVF, and OD.

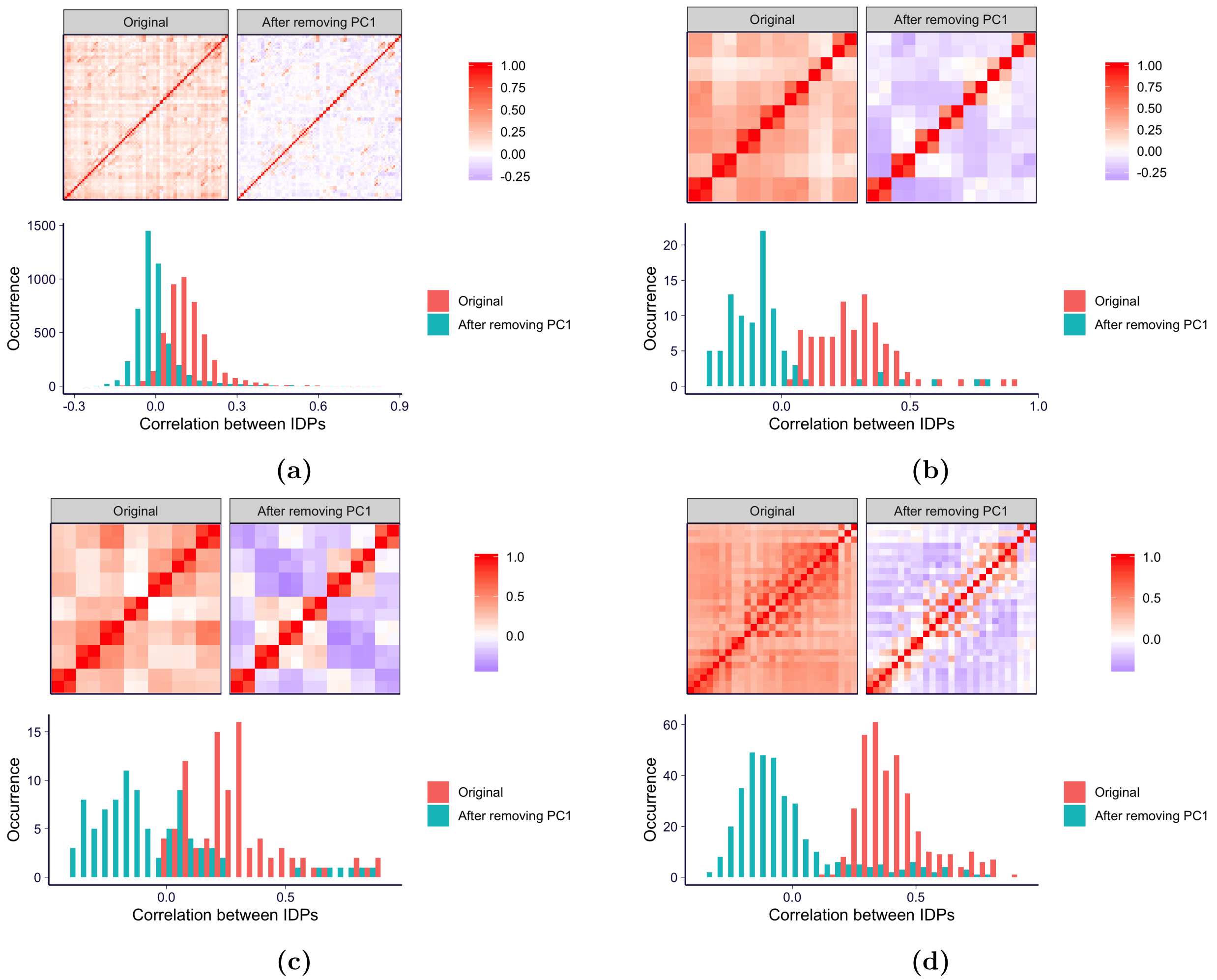

**fig. S2. The correlation between IDPs for T1 modalities** For each T1 modality, the correlation between brain IDPs are shown before and after removing PC1 by the heatmaps and the histrogram. Panel **a)** to **d)** show results for T1 modalities: gray matter volume of cortical regions, total volume of subcortical regions, gray matter volume of subcortical regions, and gray matter volume of cerebellum regions.

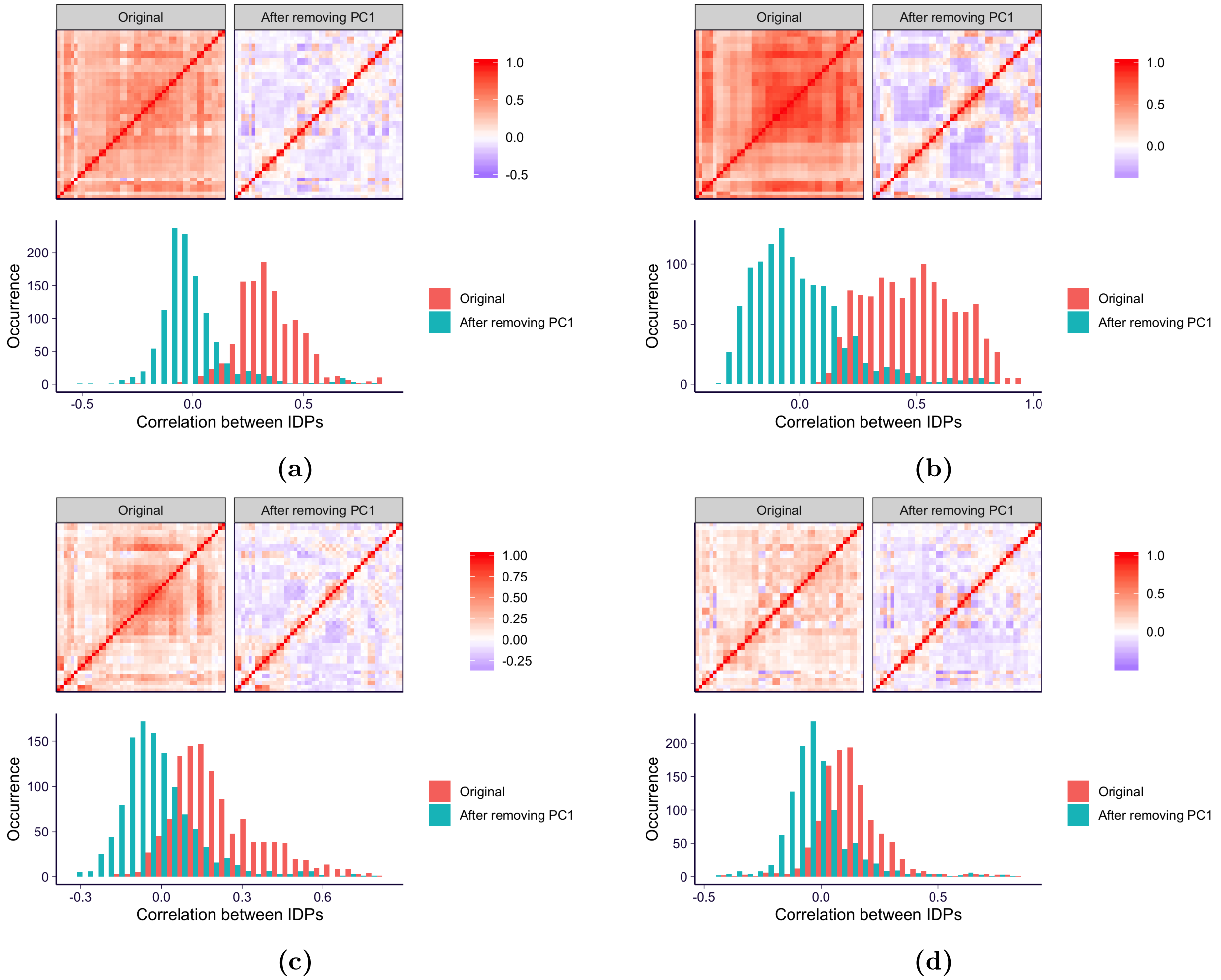

**fig. S3. The correlation between IDPs for dMRI modalities** For each dMRI modality, the correlation between brain IDPs are shown before and after removing PC1 by the heatmaps and the histrogram. Panel **a)** to **d)** show results for TBSS-based dMRI modalities: FA, ICVF, ISOVF, and OD.

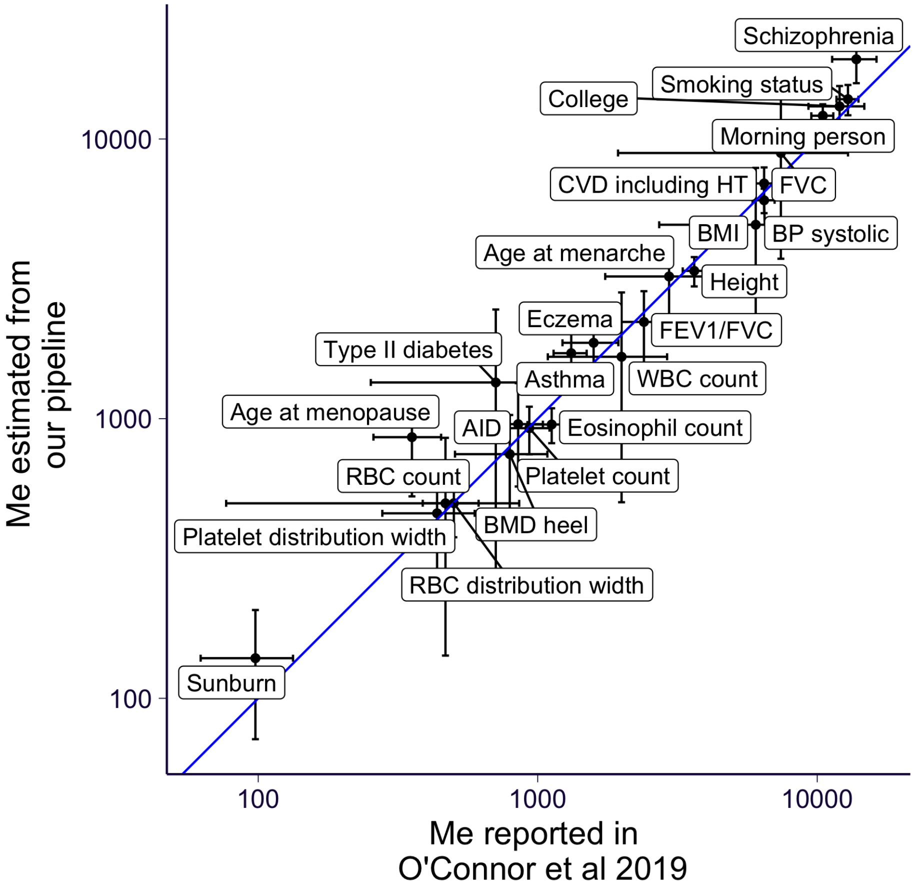

**fig. S4. Comparing estimated $M_e$ from [3] and our pipeline.** Our $M_e$ estimation pipeline is slightly different from the one being used in [3] (see more details in Methods). To check the robustness of our pipeline, we compared the estimated $M_e$ from [3] (x-axis) and our pipeline (y-axis) for 24 traits. See definition of trait abbreviations from Table 1 and Table S4 of [3]. The error bar indicates the 95% confidence interval. The blue line is the identity line ($y = x$).

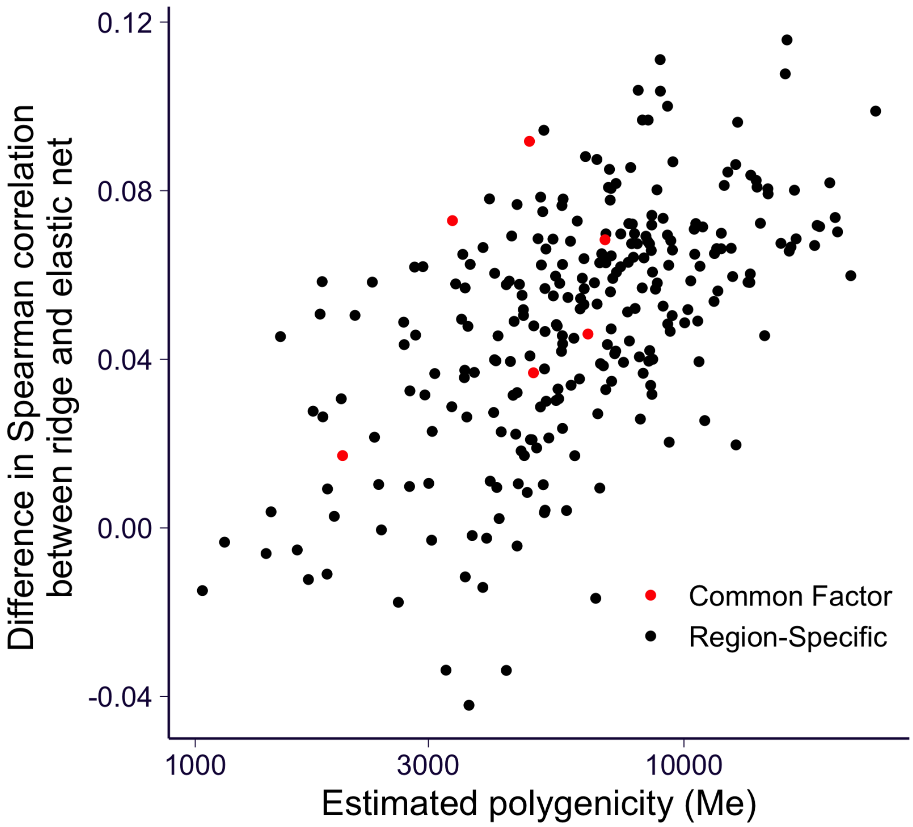

**fig. S5. Ridge predictor gain in performance vs. estimated polygenicity, $M_e$.** The x axis shows the estimated polygenicity, $M_e$, for the 522 IDPs with values significantly greater than 0 ($p < 0.05$). $M_e$ is the "the effective number of independently associated SNPs", a proxy for number of causal SNPs. The y axis shows the difference in performance between ridge predictors and elastic net predictors (in terms of the difference in Spearman correlation). The IDP PCs are in red and the rest of the brain IDPs are in black.

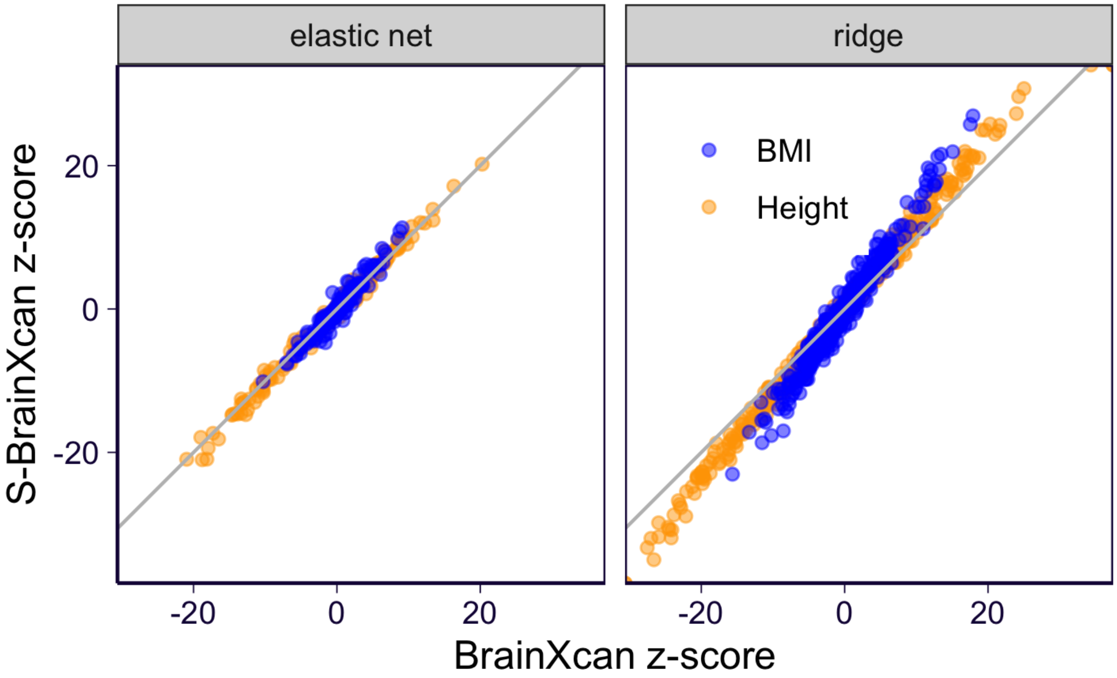

**fig. S6. Comparing individual-level BrainXcan and S-BrainXcan results on UK Biobank standing height and BMI.** We compare the BrainXcan z-scores of UK Biobank being calculated from the individual-level BrainXcan (on x-axis) and S-BrainXcan (on y-axis). For the ease of the comparison, the raw (S-)BrainXcan z-scores are shown (i.e. without permutation-based adjustment). IDP models with prediction performance greater than 0.1 (Spearman correlation) are shown.

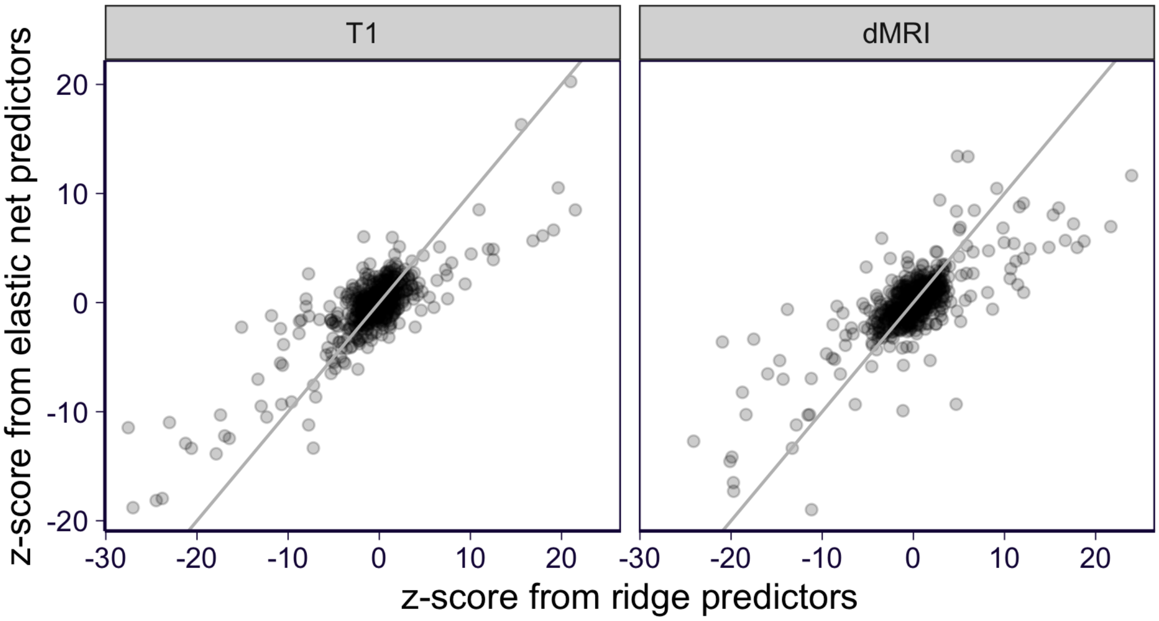

**fig. S7. Comparing the ridge and elastic net based individual-level BrainXcan results.** We compare the BrainXcan z-scores among the brain IDPs which have both ridge predictor and elastic net predictor with high quality (Spearman correlation $> 0.1$). The gray lines are the identity line ($y = x$). The permutation-based Brainxcan z-score adjustment is only applicable to S-BrainXcan. So here, the raw BrainXcan z-scores are shown.

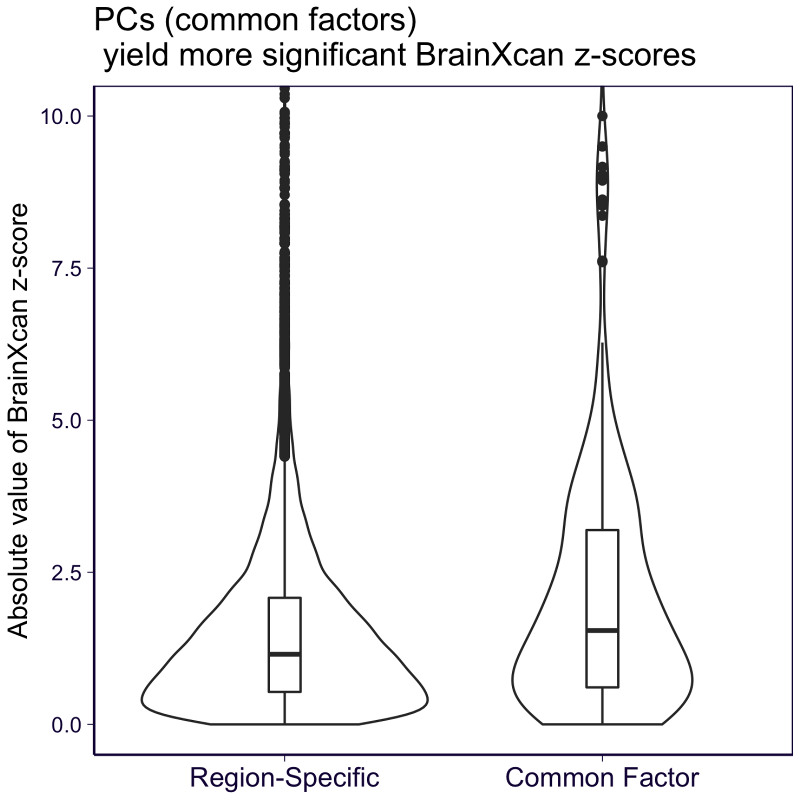

**fig. S8. Comparing the S-BrainXcan significance between region-specific IDPs and common factors.** S-BrainXcan significance is defined as the absolute value of BrainXcan z-score (with permutation-based adjustment). We compare the S-BrainXcan significance (y-axis) of region-specific IDPs and common factors (PC1 of each IDP subtype) among the brain IDPs which have ridge predictor in high quality (Spearman correlation $> 0.1$).

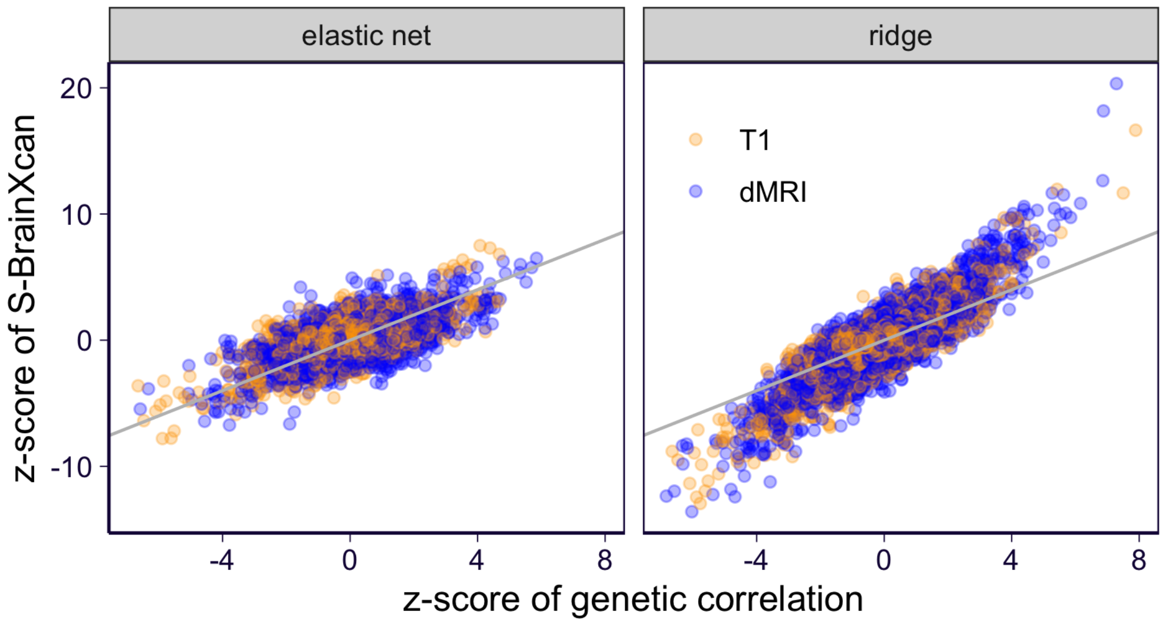

**fig. S9. Comparing z-scores of the genetic correlation and S-BrainXcan.** The z-scores of the genetic correlation (on x-axis) and the S-BrainXcan (with permutation-based adjustment; on y-axis) are shown.

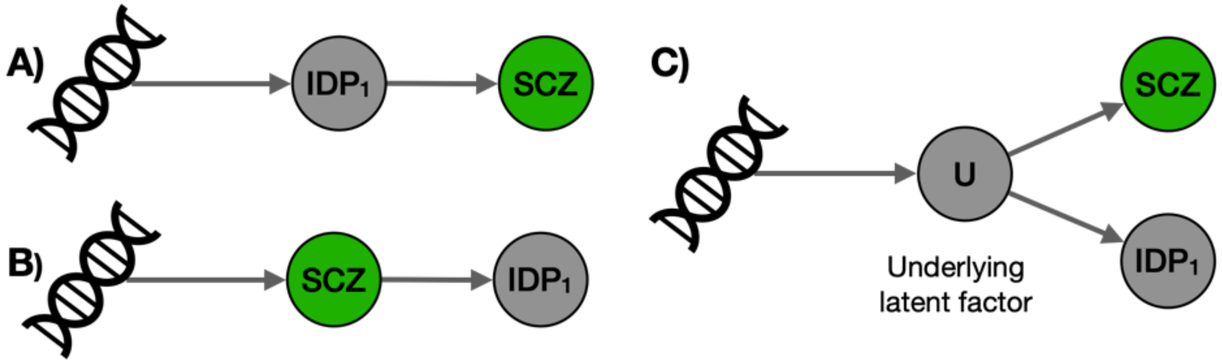

**fig. S10. Mendelizan Randomization causal flow interpretation.** Associations between brain features and the trait can arise from multiple mediating scenarios. We considered i) brain IDP alters trait, ii) trait (disease status) alters brain IDP, iii) underlying latent factor alters both trait and brain IDP. Given the power differential with current GWAS and reference image datasets, significant scenario ii) may not rule out scenario i) or iii). See discussion in text.

### Supplementary Tables

**table S1. UK Biobank brain IDPs being analyzed.** The table is available at `https://docs.google.com/spreadsheets/d/1xzLvwtIshyX1OorGnGVyDNuyDHU-zgoYhGYemf-6I10`. It contains the meta information about the brain IDPs analyzed in this paper. The columns are listed below. **ukb_field**: Field ID in UK Biobank; **modality**: indicates whether the IDP is from T1 MRI or diffusion MRI; **region**: anatomical region of the MRI measurement; **side**: indicates whether the measurement is from the left or right part of the brain; **measurement_type**: type of the measurement which also depends on the image processing procedure; **dmri_measure**: for diffusion MRI, indicate the type of statistic extracted from the measurement; **t1_anatomy_group**: for T1 MRI, indicate the anatomical grouping; **notes**: description of the IDP extracted from the UK Biobank database; **ukb_link**: link to the Field in UK Biobank website; **subtype**: IDP subtype.

**table S2. The list of 9 UK Biobank phenotypes analyzed by individual-level BrainXcan.** The table is available at `https://docs.google.com/spreadsheets/d/15OmFxAV1p_IK1l36GKjazBtOpIb5eSCOLO7JeFaTS1I`. It contains the definition of these 9 UK Biobank phenotypes. For some phenotypes, they were constructed from multiple UK Biobank fields and the aggregation method across these fields was either taking the sum ('sum') or taking logical OR ('or'). For binary phenotype, we defined its value as 1 if the field takes the desired value ('target_value'). Missing values were either treated as 0 or removed. **phenotype**: name of the phenotype; **ukb_field**: UK Biobank fields from which the phenotype was constructed; **target_value**: for binary phenotype, we defined its value as 1 if the field takes the desired value; for quantitative traits, use the value as it is (labelled as 'asis'); **aggregate_method**: for some phenotypes, they were constructed from multiple UK Biobank fields and the aggregation method across these fields was either taking the sum ('sum') or taking the logical OR ('or'); **missing_values**: missing values in a field were either treated as 0 or removed.

**table S3. The list of 35 GWAS analyzed by S-BrainXcan.** The table is available at `https://docs.google.com/spreadsheets/d/1jjDZZBsRNTvgidXXfJqXNOvFQ1U9ReeOG-WFcXtWGAg`. It contains the information about the 35 GWASs used in S-BrainXcan analysis. **phenotype**: The name of the phenotype. **phenotype_id**: The phenotype identifiers. **short_name**: The short names of the phenotypes. **sample_size**: The sample size of the GWAS. If there are different sample sizes for different SNPs, the mean sample size is shown. **portal**: The website from which the GWAS was downloaded. **filename**: The name of the raw GWAS file being downloaded.

### Supplementary Notes

### 1 Deriving bias of BrainXcan estimates

#### 1.1 A generative model of IDP/phenotype association

Recall that we consider the following generative model describing the effect of brain IDPs within a subtype on a complex trait $Y$:

$$Y = \alpha \cdot L + \sum_k \beta_k \cdot F_k + \epsilon_Y, \ \epsilon_Y \sim N(0, \sigma^2) \tag{1}$$

where $L$ represents brain-wide factor which universally affects all IDPs with the subtype. And $F_k$ represents the IDP of a specific brain region within the subtype. Eq 1 assumes that both the brain-wide factor and region-level IDPs may affect the complex trait and the effect sizes are $\alpha$ and $\beta_k$'s respectively. In the BrainXcan analysis, ideally, we are interested in both brain-wide effects ($\alpha$) and region-specific effects ($\beta_k$).

As described above, $F_k$ is partially determined by the brain-wide factor and we introduce $R_k$ to represent the rest of the variation in $F_k$ which is region-specific. Furthermore, we assume that the region-specific variations $R_k$ are independent to all other region's. More specifically, we assume

$$F_k = L + R_k, \ R_k \sim N(0, \sigma_k^2) \tag{2}$$

$$R_k \perp R_j, \ \forall k \neq j \tag{3}$$

And in this setup, the relative contribution of $L$ to $F_k$ is determined by $\sigma_k^2$.

In practice, we don't observe $F_k$. Instead, MRI imaging pipeline measures a noisy version of $F_k$. And we build genetic predictors using the noisy $F_k$ so that we manage to (partially) capture the genetically determined variation in $F_k$. In other words, let $\text{IDP}_k$ represent the predicted value of $F_k$ and we assume that

$$\text{IDP}_k = F_k + \epsilon_k, \ \epsilon_k \sim N(0, \tau_k^2) \tag{4}$$

where $\tau_k^2$ is the amount of noise when using $\text{IDP}_k$ as the proxy for $F_k$.

To further simplify the derivation, we let $\tau_j^2 = t^2, \ \forall j$ and $\sigma_j^2 = s^2, \ \forall j$. These assumptions imply equal contribution of the brain-wide factor to all regions. And they also assume that the quality of $F_k$ proxies (*i.e.* $\text{IDP}_k$) is the same across all regions. These assumptions simplify the notation and the qualitative conclusion still holds when this assumption is relaxed.

Notice that $L$ is also an unobserved latent factor. We try to capture $L$ by averaging over all $\text{IDP}_k$. In practice, we use the first principal component (PC1), which essentially is a weighted average of $\text{IDP}_k$, to approximate $L$. Using PC1 could account for the fact that the brain-wide factor does not contribute equally to all regions and predictor quality is not the same across all regions. But since here we assume equal contribution for $L$ and equal quality for $\text{IDP}_k$, we simply use unweighted average of $\text{IDP}_k$ as the proxy for $L$, which is shown below:

$$\text{PC} = \frac{1}{m} \sum_k \text{IDP}_k \tag{5}$$

$$= L + \frac{1}{m}\sum_k (R_k + \epsilon_k) \tag{6}$$

where $m$ is the number of regions within the subtype being considered.

##### 1.2 Variances and covariances among variables

Here we list the variance and covariance among model variables.

$$\text{Cov}(F_i, F_j) = \begin{cases} 1 & , i \neq j \\ 1 + s^2 & , i = j \end{cases}$$

$$\text{Cov}(\text{IDP}_i, \text{IDP}_j) = \begin{cases} 1 & , i \neq j \\ 1 + s^2 + t^2 & , i = j \end{cases} \tag{7}$$

$$\text{Var}(\text{PC}) = 1 + \frac{1}{m^2}\sum_k (s^2 + t^2) \tag{8}$$

$$\text{Cov}(\text{PC}, \text{IDP}_j) = 1 + \frac{1}{m}(s^2 + t^2) \tag{9}$$

$$\text{Cov}(L, \text{IDP}_j) = \text{Cov}(L, F_j) = 1$$

$$\text{Cov}(Y, \text{IDP}_j) = \alpha + \sum_k \beta_k + s^2 \beta_j \tag{10}$$

$$\text{Cov}(Y, \text{PC}) = \alpha + \sum_k \beta_k + \frac{1}{m}\sum s^2 \beta_k \tag{11}$$

##### 1.3 Biases of the BrainXcan associations

As described in Supplementary Notes 1.1, $\text{IDP}_k$ is a proxy of $F_k$ and PC is a proxy of $L$. In BrainXcan analysis, we fit linear regression model $Y \sim \text{IDP}_k + \text{PC}$, in which we seek to test whether there exists the region-specific effect ($\beta_k \neq 0$) so we focus on testing if the coefficient of $\text{IDP}_k$ is zero. Similarly, we also fit $Y \sim \text{PC}$ to test for a brain-wide effect of the subtype. In this section, we derive the expected value of these coefficients (coefficient of $\text{IDP}_k$ in $Y \sim \text{IDP}_k + \text{PC}$ and coefficient of PC in $Y \sim \text{PC}$) to determine how they related to the parameters of interest ($\beta_k$ and $\alpha$).

###### Coefficient of $\text{IDP}_k$

Consider fitting the linear model $Y \sim \text{IDP}_k + \text{PC}$. The coefficient of $\text{IDP}_k$ is

$$\begin{bmatrix} \text{coef IDP}_k \\ \text{coef PC} \end{bmatrix} = \begin{bmatrix} \widehat{\text{Var}}(\text{IDP}_k) & \widehat{\text{Cov}}(\text{IDP}_k, \text{PC}) \\ \widehat{\text{Cov}}(\text{IDP}_k, \text{PC}) & \widehat{\text{Var}}(\text{PC}) \end{bmatrix}^{-1} \begin{bmatrix} \widehat{\text{Cov}}(\text{IDP}_k, Y) \\ \widehat{\text{Cov}}(\text{PC}, Y) \end{bmatrix} \tag{12}$$

Taking the expected value of the coefficient, we have

$$\text{E}\left(\begin{bmatrix} \text{coef IDP}_k \\ \text{coef PC} \end{bmatrix}\right) = \begin{bmatrix} \text{Var}(\text{IDP}_k) & \text{Cov}(\text{IDP}_k, \text{PC}) \\ \text{Cov}(\text{IDP}_k, \text{PC}) & \text{Var}(\text{PC}) \end{bmatrix}^{-1} \begin{bmatrix} \text{Cov}(\text{IDP}_k, Y) \\ \text{Cov}(\text{PC}, Y) \end{bmatrix} + O_p\left(\frac{1}{n}\right) \tag{13}$$

, where $n$ is the sample size of the linear regression. And the $O_p(\cdot)$ is introduced when plugging-in variances and covariances in the places of sample variances and covariances (see more detailed discussion in [1] Appendix A).

Substituting Eq 7-11 for quantities in Eq 13, we have

$$\mathrm{E}(\text{coef IDP}_k) = \underbrace{\frac{s^2}{s^2+t^2}}_{\text{Attenuation bias}} \cdot \left[\beta_k - \underbrace{\frac{1}{m-1}\sum_{j\neq k}\beta_j}_{\text{Collider effect}}\right] + O\left(\frac{1}{n}\right) \tag{14}$$

$$= \beta_k - \frac{t^2}{s^2+t^2}\cdot\beta_k - \frac{s^2}{s^2+t^2}\sum_{j\neq k}\frac{\beta_j}{m-1} + O\left(\frac{1}{n}\right) \tag{15}$$

We note that there are two sources of bias. First, $\frac{s^2}{s^2+t^2}$ term is introduced by the fact that $\text{IDP}_k$ is a noisy version of the actual affecting variable $F_k$, which is the so called attenuation bias. Second, $\frac{1}{m-1}\sum_{j\neq k}\beta_j$ term is introduced by the fact that PC is used instead of $L$ as a covariate. As shown in Eq 6, PC captures not only $L$ but $R_k$'s which makes PC a collider variable in testing association between $Y$ and $\text{IDP}_k$.

In summary, the coefficient of $\text{IDP}_k$ in $Y \sim \text{IDP}_k + \text{PC}$ mainly captures the effect of region $k$ on $Y$ ($\beta_k$) but it also captures the average effect from all other regions. In a situation where only a few regions have non-zero effects, the second term is usually small.

**Coefficient of PC**

Consider fitting the linear model $Y \sim \text{PC}$. The coefficient of PC is

$$\text{coef PC} = \frac{\widehat{\mathrm{Cov}}(\text{PC}, Y)}{\widehat{\mathrm{Var}}(\text{PC})} \tag{16}$$

Similarly to Eq 13, to work out the expected value of PC coefficient, we plug-in variances and covariances in the places of sample variances and covariances.

$$\mathrm{E}(\text{coef PC}) = \frac{\mathrm{Cov}(\text{PC}, Y)}{\mathrm{Var}(\text{PC})} + O(\frac{1}{n}) \tag{17}$$

$$= \frac{m\alpha + (s^2+m)\sum_j \beta_j}{s^2+t^2+m} \tag{18}$$

$$= \alpha + \sum_j \beta_j + O(\frac{1}{m}) + O(\frac{1}{n}) \tag{19}$$

In summary, the coefficient of PC in $Y \sim \text{PC}$ captures the overall effect of the subtype ($\alpha + \sum_j \beta_j$).

### 2 Using IDP residual instead of fitting IDP and PC jointly

Fitting $\text{IDP}_k$ and PC jointly requires estimating the sample covariance between the predicted $\text{IDP}_k$ and PC. In summary-based BrainXcan, this estimation relies on an external LD panel, which may contain some noise or even error especially when the LD panel is not representative of the GWAS cohort. In this case, the joint model fitting is sensitive to the quality of the sample covariance estimation. To ensure the robustness of the BrainXcan test, we take an alternative approach which avoid estimating the sample covariance.

In the alternative approach, we fit $Y \sim \text{resIDP}_k$ instead where $\text{resIDP}_k$ is the predicted value of $\text{IDP}_k$ residual (after regressing out PC). In this section, we show that the coefficient of $\text{IDP}_k$ in the joint model $Y \sim \text{IDP}_k + \text{PC}$ is approximately equivalent to the result of a two-step approach:

1. Regress out PC from $\text{IDP}_k$ and keep the residual $r_k$.
2. Obtain coefficient of $r_k$ in $Y \sim r_k$.

First of all, we can calculate $r_k$ from model $\text{IDP}_k \sim \text{PC}$.

$$r_k = \text{IDP}_k - a_k \cdot \text{PC} \tag{20}$$

$$a_k = \frac{\widehat{\text{Cov}}(\text{IDP}_k, \text{PC})}{\widehat{\text{Var}}(\text{PC})} \tag{21}$$

So, we have

$$\text{Cov}(Y, r_k) = \text{Cov}(Y, \text{IDP}_k) - a_k \cdot \text{Cov}(Y, \text{PC}) \tag{22}$$

$$\text{Var}(r_k) = \text{Var}(\text{IDP}_k) - 2 \cdot a_k \cdot \text{Cov}(\text{IDP}_k, \text{PC}) + a_k^2 \cdot \text{Var}(\text{PC}) \tag{23}$$

And in step 2, we can obtain the coefficient of $r_k$ from $Y \sim r_k$.

$$\text{coef } r_k = \frac{\widehat{\text{Cov}}(Y, r_k)}{\widehat{\text{Var}}(r_k)} \tag{24}$$

$$\text{E}(\text{coef } r_k) \approx \frac{\text{Cov}(Y, r_k)}{\text{Var}(r_k)} \tag{25}$$

$$= \frac{s^2}{s^2 + t^2} \cdot \left[\beta_k - \frac{1}{m-1} \sum_{j \neq k} \beta_j\right] \tag{26}$$

Comparing Eq 14 and 26, we can conclude that $\text{E}(\text{coef}\, \text{IDP}_k) \approx \text{E}(\text{coef}\, r_k)$. And this result indicates that regressing the outcome $Y$ on $\text{IDP}_k$ and PC jointly is approximately equivalent to regressing $Y$ on the residual of $\text{IDP}_k$.

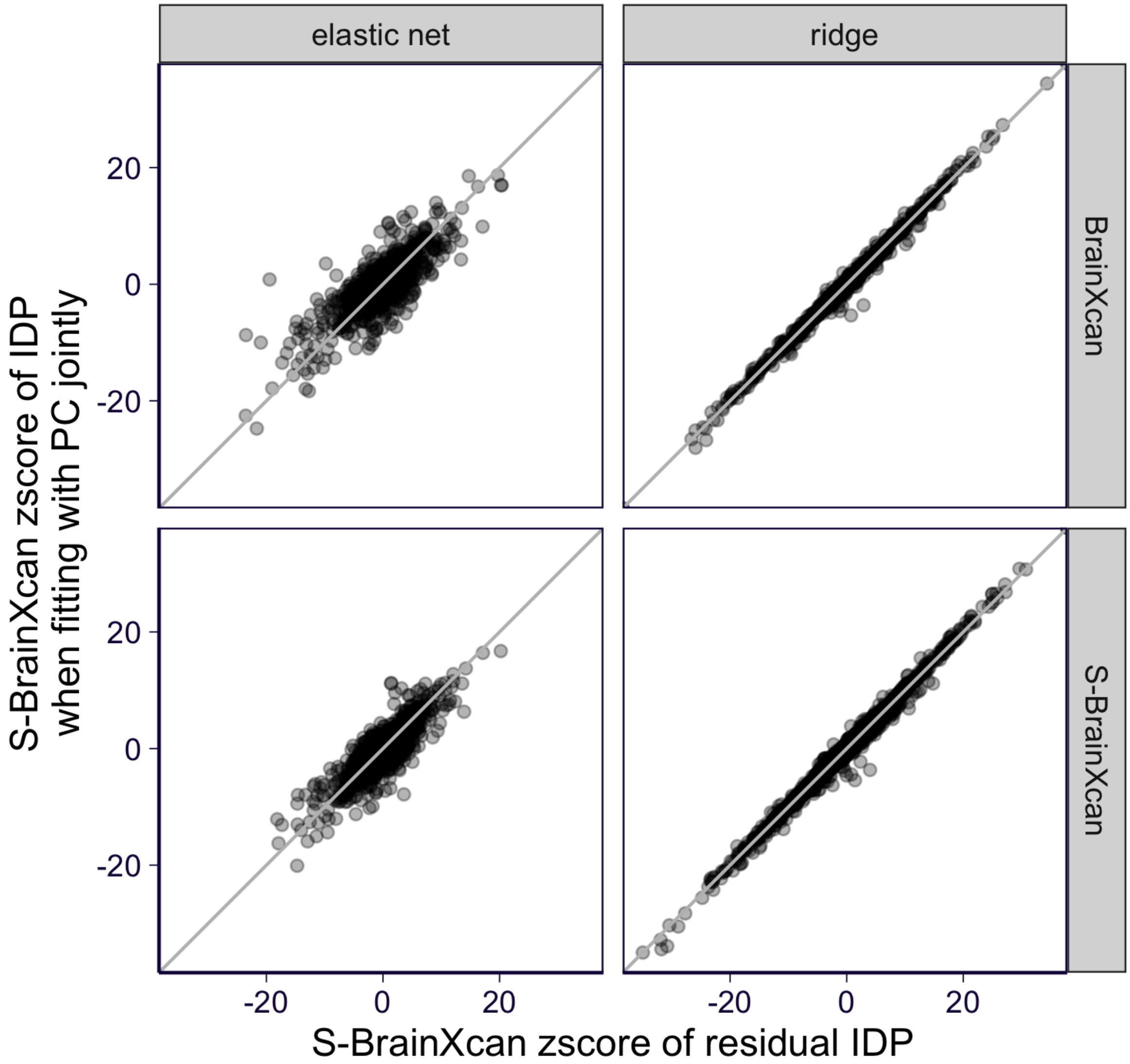

**Additional fig. A1. Comparing BrainXcan results from residual IDP and IDP adjusted by PC.** We compare the BrainXcan z-scores obtained from $Y \sim$ resIDP (x-axis) and $Y \sim$ IDP+PC (y-axis). The top row shows results from individual-level BrainXcan and the bottom row shows results from summary BrainXcan. For the ease of the comparison, the raw (S-)BrainXcan z-scores are shown (i.e. without permutation-based adjustment). IDP models with prediction performance greater than 0.1 (Spearman correlation) are shown. The gray lines are the identity line ($y = x$). All GWASs are shown.

In practice, instead of calculating the residual $\text{IDP}_k$ by regressing out predicted PC from predicted $\text{IDP}_k$ (which still relies on covariance between PC and $\text{IDP}_k$), we build the genetic predictor of $\text{IDP}_k$ residual and predict the residual $\text{IDP}_k$ (which is called $\text{resIDP}_k$) directly. We show empirically that the coefficient from $Y \sim \text{resIDP}_k$ is similar to coef $\text{IDP}_k$ (fig. A1).

#### 3 Aggregating Mendelian Randomization test results by extending the Aggregated Cauchy Association test (ACAT) method

The ACAT method [2] is a "meta-analysis" approach that leverages the fact that averages of possible correlated Cauchy random variables are Cauchy-distributed [4]. The method ignores the direction of the association since it uses p-values alone. However, it is obvious that two studies with opposite direction of effects should somewhat cancel each other and yield a less significant p-value when combined. Here, we propose an approach to take the sign into account.

ACAT combines potentially dependent p-values as follows:

$$T = \sum_i \tan[(\frac{1}{2} - p_i) \cdot \pi] \tag{27}$$

$$p_{\text{ACAT}} = \frac{1}{2} - \frac{\arctan(T/N)}{\pi} \tag{28}$$

where $N$ is the total number of p-values being combined and $T$ is the test statistic which follows Cauchy distribution under the null. So, we want to apply ACAT to combine the results of multiple Mendelian Randomization tests.

In the original use case of the ACAT method, p-values are combined without considering the direction of the effect. In our specific example, various Mendelian Randomization tests yield p-values ($p_i$) and direction of the effects ($s_i$). So, we want to construct a variation of ACAT such that the direction of the effect is considered. Specifically, to combine p-values $p_1, \cdots, p_N$ with signs $s_1, \cdots, s_N$, if we assume the "+1" direction is the direction of true signal, we take the signs into consideration by modifying Eq 27:

$$T_{+1} = \sum_i \{s_i \cdot \tan[(\frac{1}{2} - p_i) \cdot \pi]\}, \tag{29}$$

or for the negative direction

$$T_{\text{-}1} = -\sum_i \{s_i \cdot \tan[(\frac{1}{2} - p_i) \cdot \pi]\}. \tag{30}$$

More generally, considering $s \in \{-1, +1\}$ as the true direction:

$$T_s = s \cdot \sum_i \{s_i \cdot \tan[(\frac{1}{2} - p_i) \cdot \pi]\}. \tag{31}$$

In practice, both "+1" and "-1" directions are possible so we should test both and combine the two p-values at the end. For this purpose, we propose the following meta-analysis approach, signed ACAT (SACAT), which takes p-values $p_1, \cdots, p_N$ with signs $s_1, \cdots, s_N$ and return the combined p-value and sign:

$$p_s = \frac{1}{2} - \frac{\arctan(T_s/N)}{\pi} \tag{32}$$

$$p_{\mathrm{SACAT}} = 2 \cdot \min_s p_s \tag{33}$$

$$s_{\mathrm{SACAT}} = \arg\min_s p_s \tag{34}$$

The $p_{\mathrm{SACAT}}$ is derived using the fact that $T_{+1} = -T_{\text{-}1}$ implies $p_+ + p_- = 1$ and therefore $P\{\min(p_+, p_-) < u\} = P\{\min(p_+, (1 - p_+)) < u\} = P\{\min(p_-, (1 - p_-)) < u\} = 2u$ with $u$ taking values between 0 and 1/2.

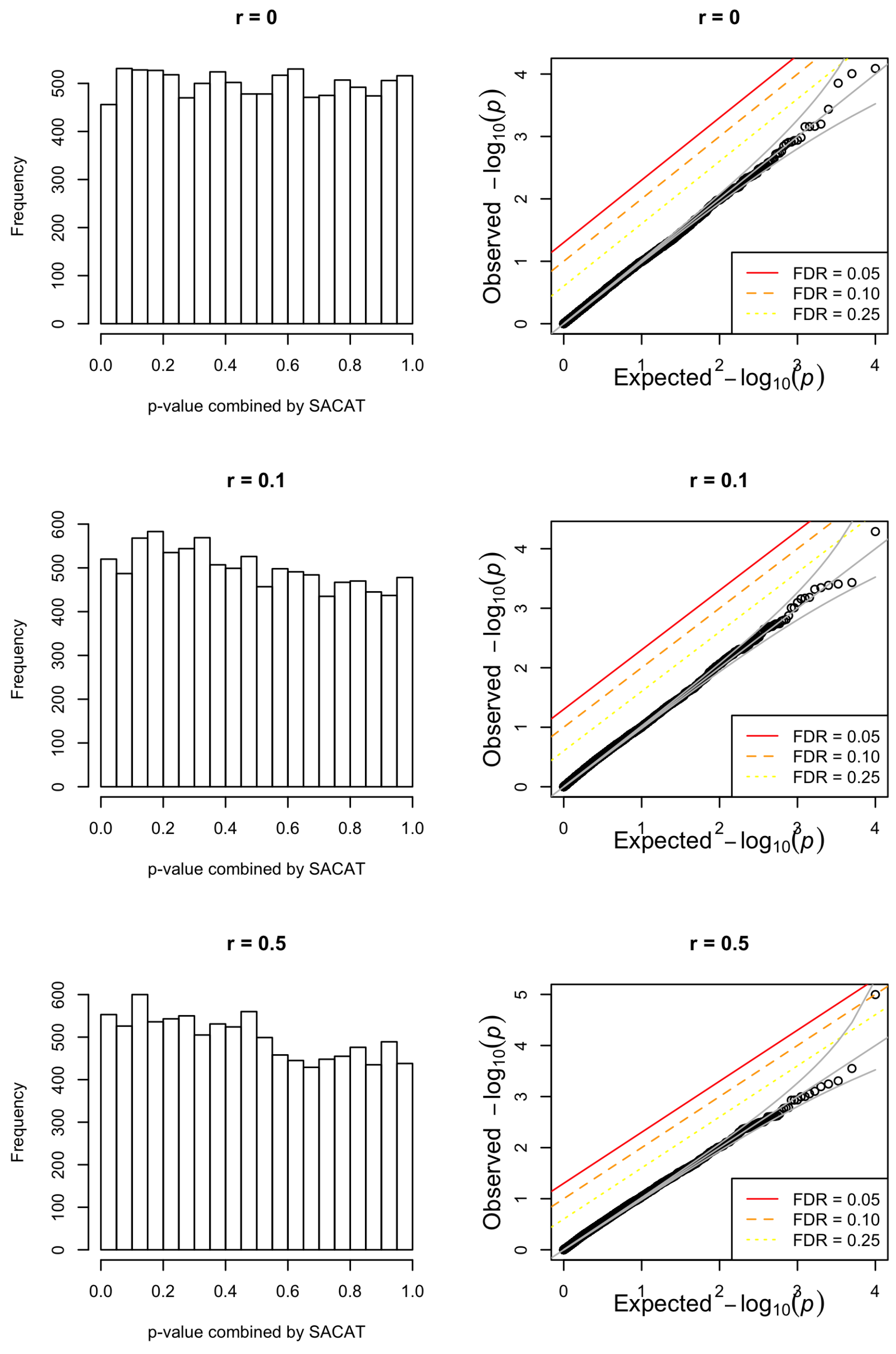

**Additional fig. A2. SACAT based p-value distribution under the global null.** The simulation results of signed-ACAT (SACAT) are shown. The distribution of the SACAT-based meta-analyzed p-values are shown with histogram and QQ-plot (against uniform distribution). Each row shows the results from simulating the global null with dependency structure controlled by $r$. As $r$ increases, the p-values being combined are more dependent and when $r = 0$, the p-values being combined are independent. These figures show that the SACAT is relatively well calibrated for different values of correlation between entries. See more simulation details in Supplementary Notes 3.

To examine the calibration of SACAT, we perform a simulation study in which we simulate from the global null using the procedure in below:

1. For $i = 1, \cdots, 20$, we simulate $Z_i \sim N(0, 1)$ where $\text{Cov}(Z_i, Z_j) = r$ except $\text{Cov}(Z_1, Z_j) = -r$.
2. Calculate $p_i = 2 \times \Pr(|Z| > Z_i; Z \sim N(0, 1))$.
3. Repeat the above two steps 10000 times for $r = 0, 0.1, 0.5$ respectively.

The dependence among p-values is induced by $r \neq 0$. Fig. A2 shows that the proposed SACAT is well calibrated under the global null since the resulting meta-analyzed p-values are roughly uniformly distributed and the points in the qq-plot against the null distribution lie on the identity line for all three values of dependence $r$.

### 4 Caveats on interpreting Mendelian randomization results

There are two caveats that need to be considered when interpreting Mendelian randomization results. One is that we first select the IDP-trait pair based on their association. Therefore, the p-values of the Mendelian randomization will not be well-calibrated, i.e. even under the null of no causal link, the p-value will be biased towards smaller values. Given this bias, we should not use the Mendelian Randomization p-values to claim significance but as scores to discern between possible direction of the causal flow.

The second caveat relates to the power difference between reference image and GWAS studies. Currently, reference image data have much smaller sample sizes (n $\sim$ 30K) compared to GWAS studies of complex traits (n $\sim$ 100K to 1M). We consider three main mediating scenarios depicted in fig. S10. In scenario i) the brain feature mediates the genetic association with the phenotype, i.e. genetic risk factors alter the brain feature which in turn alters the risk for the phenotype. In scenario ii) genetic factors affect the phenotype which in turn alter the brain feature. In scenario iii) genetic factors affect an underlying latent factor which alter both the phenotype and the brain feature.

A significant i) and not significant ii) can be interpreted as evidence that the brain feature alteration is affecting the phenotype given the higher power of GWAS studies in general. However, a significant scenario ii) and non significant scenario i) could simply mean that the instruments (strongly associated SNPs and their effect sizes) for the brain feature are not reliable enough to yield significance. In this case, scenario ii) should be considered supported by the data but scenarios i) and iii) should not be ruled out.
